# RESEARCH PROTOCOL: Large-scale evidence generation and evaluation across a network of databases for type 2 diabetes mellitus

**DOI:** 10.1101/2021.09.27.21264139

**Authors:** Rohan Khera, Martijn J Scheumie, Yuan Lu, Anna Ostropolets, Ruijun Chen, George Hripcsak, Patrick B Ryan, Harlan M Krumholz, Marc A Suchard

## Abstract

**Background:** Therapeutic options for type 2 diabetes mellitus (T2DM) have expanded over the last decade with the emergence of sodium-glucose co-transporter-2 (SGLT2) inhibitors and glucagon-like peptide-1 (GLP1) receptor agonists, which reduced the risk of major cardiovascular events in randomized controlled trials (RCTs). Cardiovascular evidence for older second-line agents, such as sulfonylureas, and direct head-to-head comparisons, including with dipeptidyl peptidase 4 (DPP4) inhibitors, are lacking, leaving a critical gap in our understanding of the relative effects of T2DM agents on cardiovascular risk and on patient-centered safety outcomes.

**Methods and Analysis:** The Large-Scale Evidence Generations Across a Network of Databases for T2DM (LEGEND-T2DM) initiative is a series of systematic, large-scale, multinational, real-world comparative cardiovascular effectiveness and safety studies of all 4 major second-line anti-hyperglycemic agents including SGLT2 inhibitor, GLP1 receptor agonist, DPP4 inhibitor and sulfonylureas. LEGEND-T2DM will leverage the Observational Health Data Science and Informatics (OHDSI) community that provides access to a global network of administrative claims and electronic health record (EHR) data sources. Committed data partners represent 190 million patients in the US and about 50 million internationally. LEGEND-T2DM will identify all adult, T2DM patients who newly initiate a traditionally second-line T2DM agent, including individuals with and without established cardiovascular disease. Using an active comparator, new-user cohort design, LEGEND-T2DM will execute all pairwise class-vs-class and drug-vs-drug comparisons in each data source that meet a minimum patient count of 1,000 per arm and extensive study diagnostics that assess reliability and generalizability through cohort balance and equipoise to examine the relative risk of cardiovascular and safety outcomes. The primary cardiovascular outcomes include a 3-point and a 4-point composite of major adverse cardiovascular events, and series of safety outcomes. The study will pursue data-driven, large-scale propensity adjustment for measured confounding, a large set of negative control outcome experiments to address unmeasured and systematic bias.

**Ethics and Dissemination:** The study ensures data safety through a federated analytic approach and follows research best practices, including prespecification and full disclosure of hypotheses tested and their results. LEGEND-T2DM is dedicated to open science and transparency and will publicly share all our analytic code from reproducible cohort definitions through turn-key software, enabling other research groups to leverage our methods, data, and results in order to verify and extend our findings.

## 1 Rationale and Background

The landscape of therapeutic options for type 2 diabetes mellitus (T2DM) has been dramatically transformed over the last decade [1]. The emergence of drugs targeting the sodium-glucose co-transporter-2 (SGLT2) and the glucagon-like peptide-1 (GLP1) receptor has expanded the role of T2DM agents from lowering blood glucose to directly reducing cardiovascular risk [2]. A series of large randomized clinical trials designed to evaluate the cardiovascular safety of SGLT2 inhibitors and GLP1 receptor agonists found that use of many of these agents led to a reduction in major adverse cardiovascular events, including myocardial infarction, hospitalization for heart failure, and cardiovascular mortality [3–6]. However, other T2DM drugs widely used before the introduction of these novel agents, such as sulfonylureas, did not undergo similarly comprehensive trials to evaluate their cardiovascular efficacy or safety. Moreover, direct comparisons of newer agents with dipeptidyl peptidase-4 (DPP4) inhibitors, with neutral effects on major cardiovascular outcomes [7–10], have not been conducted. Nevertheless, DPP4 inhibitors and sulfonylureas continue to be used in clinical practice and are recommended as second-line T2DM agents in national clinical practice guidelines.

Several challenges remain in formulating T2DM treatment recommendations based on existing evidence [11]. First, trials of novel agents did not pursue head-to-head comparisons to older agents and were instead designed as additive treatments on the background of commonly used T2DM agents. Therefore, the relative cardiovascular efficacy and safety of novel compared with older agents is not known, and indirect estimates have relied on summary-level data restricted to common comparators [12–14] and are less reliable [15, 16]. Second, trials of novel agents have tested individual drugs against placebo, but have not directly compared SGLT2 inhibitors with GLP1 receptor agonists in reducing adverse cardiovascular event risk. Moreover, there is no evidence to guide the use of individual drugs within each class and across different drug classes, particularly among patients at lower cardiovascular risk than recruited in clinical trials. Third, randomized trials focused on cardiovascular efficacy and safety, but were not powered to adequately assess the safety of these agents across a spectrum of non-cardiovascular outcomes. Finally, restricted enrollment across regions, and subgroups of age, sex, and race further limits the efficacy and safety assessment that may guide individual patients’ treatment.

Evidence gaps from these trials also pose a challenge in designing treatment algorithms, which rely on comparative effectiveness and safety of drugs. Perhaps, as a result, there is large variation in clinical practice guidelines and in clinical practice with regard to these medications, with many patients initiated on the newer therapies and many others treated with older regimens [17–21]. Among the second-line options, there is much variation with respect to the order of drugs used. This lack of consensus about the best approach provides an opportunity for systematic, large-scale observational studies.

## 2 Study Objectives

To inform critical decisions facing patients with diabetes, their caregivers, clinicians, policymakers and healthcare system leaders, we have launched the Large-Scale Evidence Generation and Evaluation across a Network of Databases for Diabetes (LEGEND-T2DM) initiative to execute a series of comprehensive observational studies to compare cardiovascular outcome rates and safety of second-line T2DM glucose-lowering agents. Specifically, these studies aim

1. To determine, through systematic evaluation, the comparative effectiveness of traditionally second-line T2DM agents, SGLT2 inhibitors and GLP1 receptor agonists, with each other and with DPP4 inhibitors and sulfonylureas, for cardiovascular outcomes.
2. To determine, through systematic evaluation, the comparative safety of traditionally second-line T2DM agents among patients with T2DM.
3. To assess heterogeneity in effectiveness and safety of traditionally second-line T2DM agents among key patient subgroups: Using stratified patient cohorts, we will quantify differential effectiveness and safety across subgroups of patients based on age, sex, race, renal impairment, and baseline cardiovascular risk.

## 3 Research Methods

LEGEND-T2DM will execute three systematic, large-scale observational studies of second-line T2DM agents to estimate the relative risks of cardiovascular effectiveness and safety outcomes.

1. The **Class-vs-Class Study** will provide all pairwise comparisons between the four major T2DM agent classes to evaluate their comparative effects on cardiovascular risk (Objective 1) and patient-centered safety outcomes (Objective 2);
2. The **Drug-vs-Drug Study** will furnish head-to-head pairwise comparisons between individual agents within and across classes (both Objectives 1 and 2); and The **Heterogeneity Study** will refine these comparisons for T2DM patients for important subgroups (Objective 3). In contrast to a single comparison approach, LEGEND-T2DM will provide a comprehensive view of the findings and their consistency across populations, drugs, and outcomes. We will model each study on our successful collaborative research evaluating the comparative effectiveness of antihypertensives recently published in *The Lancet* [22].

Table 1 list the four major T2DM agent classes and the individual agents licensed in the U_5_.S_+_._6+_w_4_it_+_h_7_in each class. We will examine all 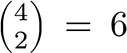 class-wise comparisons and all 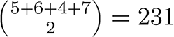 ingredient-wise comparisons.

**Table 1:**
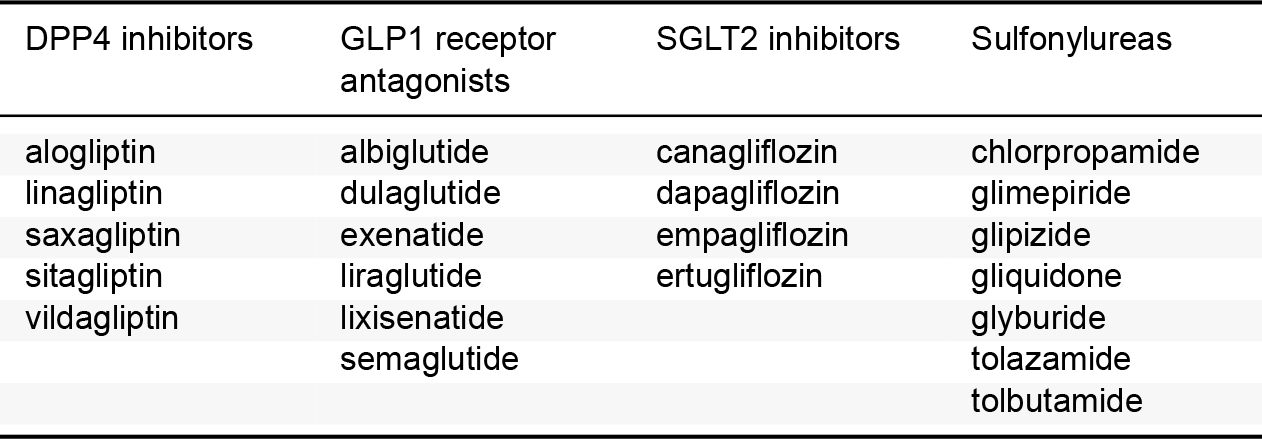
T2DM drug classes and individual agents within each class

| DPP4 inhibitors | GLP1 receptor antagonists | SGLT2 inhibitors | Sulfonylureas |
| --- | --- | --- | --- |
| alogliptin | albiglutide | canagliflozin | chlorpropamide |
| linagliptin | dulaglutide | dapagliflozin | glimepiride |
| saxagliptin | exenatide | empagliflozin | glipizide |
| sitagliptin | liraglutide | ertugliflozin | gliquidone |
| vildagliptin | lixisenatide |  | glyburide |
|  | semaglutide |  | tolazamide |
|  |  |  | tolbutamide |

**Table 2:** Committed LEGEND-T2DM data sources and the populations they cover.

| Data source | Population | Patients | History | Data capture process and short description |
| --- | --- | --- | --- | --- |
| **Administrative claims** |  |  |  |  |
| IBM MarketScan Commercial Claims and Encounters (CCAE) | Commercially insured, < 65 years | 142M | 2000 – | Adjudicated health insurance claims (e.g. inpatient, outpatient, and outpatient pharmacy) from large employers and health plans who provide private healthcare coverage to employees, their spouses and dependents. |
| IBM MarketScan Medicare Supplemental Database (MDCR) | Commercially insured, 65+ years | 10M | 2000 – | Adjudicated health insurance claims of retirees with primary or Medicare supplemental coverage through privately insured fee-for-service, point-of-service or capitated health plans. |
| IBM MarketScan Multi-State Medicaid Database (MDCD) | Medicaid enrollees, racially diverse | 26M | 2006 – | Adjudicated health insurance claims for Medicaid enrollees from multiple states and includes hospital discharge diagnoses, outpatient diagnoses and procedures, and outpatient pharmacy claims. |
| IQVIA Open Claims (IOC) | General | 160M | 2010 – | Pre-adjudicated claims at the anonymized patient-level collected from office-based physicians and specialists via office management software and clearinghouse switch sources for the purpose of reimbursement. |
| Japan Medical Data Center (JMDC) | Japan, general | 5.5M | 2005 – | Data from 60 society-managed health insurance plans covering workers aged 18 to 65 and their dependents. |
| Korea National Health Insurance Service (NHIS) | 2% random sample of South Korea | 1M | 2002 – | National administrative claims database covering the South Korean population. |
| Optum Clinformatics Data Mart (Optum) | Commercially or Medicare insured | 85M | 2000 – | Inpatient and outpatient healthcare insurance claims. |
| **Electronic health records (EHRs)** |  |  |  |  |
| Columbia University Irving Medical Center (CIUMC) | Academic medical center patients, racially diverse | 6M | 1989 – | General practice, specialists and inpatient hospital services from the New York-Presbyterian hospital and affiliated academic physician practices in New York. |
| Department of Veterans Affairs (VA) | Veterans, older, racially diverse | 12M | 2000 – | National VA health care system, the largest integrated provider of medical services in the US, provided at 170 VA medical centers and 1,063 outpatient sites. |
| Information System for Research in Primary Care (SIDIAP) | 80% of all Catalonia (Spain) | 7.7M | 2006 – | Primary care partially linked to inpatient data with pharmacy dispensations and primary care laboratories. Healthcare is universal and taxpayer funded in the region, and PCPs are gatekeepers for all care and responsible for repeat prescriptions. |
| IQVIA Disease Analyzer Germany (DAG) | Germany, general | 37M | 1992 – | Collection from patient management software used by general practitioners and selected specialists to document patients' medical records within their office-based practice during a visit. |
| Optum Electronic Health Records (OptumEHR) | US, general | 93M | 2006 – | Clinical information, prescriptions, lab results, vital signs, body measurements, diagnoses and procedures derived from clinical notes using natural language processing. |
| Yale New Haven Health System (YNHHS) | Academic medical center patients | 2M | 2013 – | General practice, specialists and inpatient hospital services from the YNHHS in Connecticut. |

For each comparison, we are interested in the relative risk of each of the cardiovascular and safety outcomes listed in Table 3.

**Table 3:** LEGEND-T2DM study outcomes

| Phenotype | Brief logical description | Prior development |
| --- | --- | --- |
| **Cardiovascular outcomes** |  |  |
| 3-point MACE | Condition record of acute myocardial infarction, hemorrhagic or ischemic stroke or sudden cardiac death during an inpatient or ER visit | [49–61] |
| 4-point MACE | 3-Point MACE + inpatient or ER visit (hospitalization) with heart failure condition record | [44,49–67] |
| Acute myocardial infarction | Condition record of acute myocardial infarction during an inpatient or ER visit | [49–54] |
| Acute renal failure | Condition record of acute renal failure during an inpatient or ER visit | [47,68–75] |
| Glycemic control | First hemoglobin A1c measurement with value $\leq 7\%$ | [76] |
| Hospitalization with heart failure | Inpatient or ER visit with heart failure condition record | [44,62–67] |
| Measured renal dysfunction | First creatinine measurement with value $> 3$ mg/dL | [75] |
| Revascularization | Procedure record of percutaneous coronary intervention or coronary artery bypass grafting during an inpatient or ER visit | [45] |
| Stroke | Condition record of hemorrhagic or ischemic stroke during an inpatient or ER visit | [55–60] |
| Sudden cardiac death | Condition record of sudden cardiac death during an inpatient or ER visit | [52,61] |
| **Patient-centered safety outcomes** |  |  |
| Abnormal weight gain | Abnormal weight gain record of any type; successive records with $> 90$ day gap are considered independent episodes; note, weight measurements not used | [77] |
| Abnormal weight loss | Abnormal weight loss record of any type; successive records with $> 90$ day gap are considered independent episodes; note, weight measurements not used | [78] |
| Acute pancreatitis | Condition record of acute pancreatitis during an inpatient or ER visit | [79–82] |
| All-cause mortality | Death record of any type | [52,83,84] |
| Bladder cancer | Malignant tumor of urinary bladder condition record of any type; limited to earliest event per person |  |
| Bone fracture | Bone fracture condition record of any type; successive records with $> 90$ day gap are considered independent episodes | |
| Breast cancer | Malignant tumor of breast condition record of any type; limited to earliest event per person |  |
| Diabetic ketoacidosis | Diabetic ketoacidosis condition record during an inpatient or ER visit | [46,85] |
| Diarrhea | Diarrhea condition record of any type; successive records with $> 30$ day gap are considered independent episodes | [86–88] |
| Genitourinary infection | Condition record of any type of genital or urinary tract infection during an outpatient or ER visit | [89] |

Table 3: LEGEND-T2DM study outcomes (*continued*)
| Phenotype | Brief logical description | Prior development |
| --- | --- | --- |
| Hyperkalemia | Condition record for hyperkalemia or potassium measurements > 5.6 mmol/L; successive records with >90 day gap are considered independent episodes | [90–92] |
| Hypoglycemia | Hypoglycemia condition record of any type; successive records with > 90 day gap are considered independent episodes | [93] |
| Hypotension | Hypotension condition record of any type; successive records with > 90 day gap are considered independent episodes | [94] |
| Joint pain | Joint pain condition record of any type; successive records with > 90 days gap are considered independent episodes |  |
| Lower extremity amputation | Procedure record of below knee lower extremity amputation during inpatient or outpatient visit | [44,48] |
| Nausea | Nausea condition record of any type; successive records with > 30 day gap are considered independent episodes | [95–97] |
| Peripheral edema | Edema condition record of any type; successive records with > 180 day gap are considered independent episodes |  |
| Photosensitivity | Condition record of drug-induced photosensitivity during any type of visit |  |
| Renal cancer | Primary malignant neoplasm of kidney condition record of any type; limited to earliest event per person |  |
| Thyroid tumor | Neoplasm of thyroid gland condition record of any type; limited to earliest event per person |  |
| Venous thromboembolism | Venous thromboembolism condition record of any type; successive records with > 180 day gap are considered independent episodes | [98–101] |
| Vomiting | Vomiting condition record of any type; successive records with > 30 day gap are considered independent episodes | [95–97] |

### 3.1 Study Design

For each study, we will employ an active comparator, new-user cohort design [23–25]. New-user cohort design is advocated as the primary design to be considered for comparative effectiveness and drug safety [26–28]. By identifying patients who start a new treatment course and using therapy initiation as the start of follow-up, the new-user design models an randomized controlled trial (RCT) where treatment commences at the index study visit. Exploiting such an index date allows a clear separation of baseline patient characteristics that occur prior to index date and are usable as covariates in the analysis without concern of inadvertently introducing mediator variables that arise between exposure and outcome [29]. Excluding prevalent users as those without a sufficient washout period prior to first exposure occurrence further reduces bias due to balancing mediators on the causal pathway, time-varying hazards, and depletion of susceptibles [28, 30]. Our systematic framework across studies further will address residual confounding, publication bias, and p-hacking using data-driven, large-scale propensity adjustment for measured confounding [31], a large set of negative control outcome experiments to address unmeasured and systematic bias [32–34], and full disclosure of hypotheses tested [35]. Figure 1 illustrates our design for all studies that the following sections describe in more detail.

**Figure 1:**
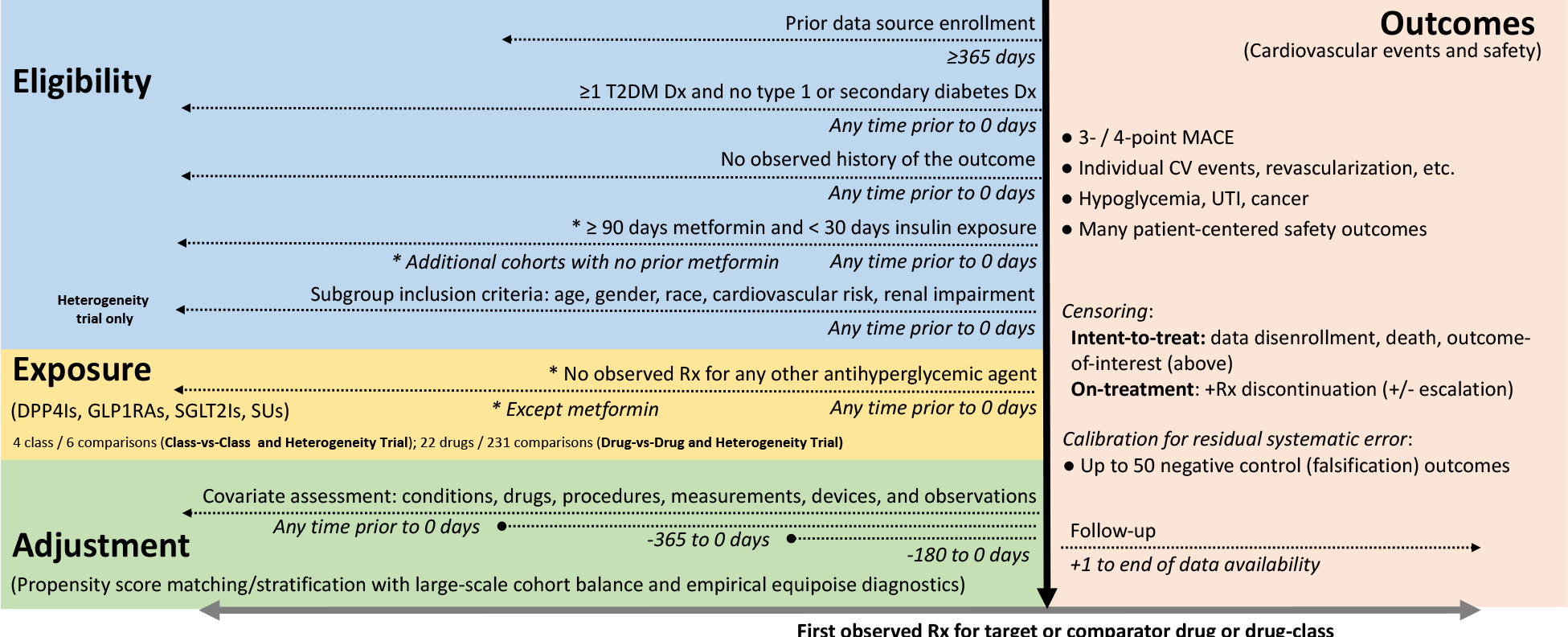
Schematic of LEGEND-T2DM new-user cohort design for the Class-vs-Class, Drug-vs-Drug and Heterogeneity studies.

### 3.2 Data Sources

We will execute LEGEND-T2DM as a series of OHDSI network studies. All data partners within OHDSI are encouraged to participate voluntarily and can do so conveniently, because of the community’s shared Observational Medical Outcomes Partnership (OMOP) common data model (CDM) and OHDSI tool-stack. Many OHDSI community data partners have already committed to participate and we will recruit further data partners through OHDSI’s standard recruitment process, which includes protocol publication on OHDSI’s GitHub, an announcement in OHDSI’s research forum, presentation at the weekly OHDSI all-hands-on meeting and direct requests to data holders.

Table 2 lists the 13 already committed data sources for LEGEND-T2DM; these sources encompass a large variety of practice types and populations. For each data source, we report a brief description and size of the population it represents and its patient capture process and start date. While the earliest patient capture begins in 1989 (CUIMC), the vast majority come from the mid-2000s to today, providing almost two decades of T2DM treatment coverage. US populations include those commercially and publicly insured, enriched for older individuals (MDCR, VA), lower socioeconomic status (MDCD), and racially diverse (VA >20% Black or African American, CUIMC 8%). The US data sources may capture the same patients across multiple sources. Different views of the same patients are an advantage in capturing the diversity of real-world health events that patients experience. Across CCAE (commercially insured), MCDR (Medicare) and MCDC (Medicaid), we expect little overlap in terms of the same observations recorded at the same time for a patient; patients can flow between sources (e.g., a CCAE patient who retires can opt-in to become an MDCR patient), but the enrollment time periods stand distinct. On the other hand, Op-tum, PanTher, OpenClaims, CUIMC and YNHHS may overlap in time with the other US data sources. While it remains against licensing agreements to attempt to link patients between most data sources, Optum reports <20% overlap between their claims and EHR data sources that is reassuringly small. All data sources will receive institutional review board approval or exemption for their participation before executing LEGEND-T2DM.

### 3.3 Study Population

We will include all subjects in a data source who meet inclusion criteria for one or more traditionally second-line T2DM agent exposure cohorts. Broadly, these cohorts will consist of T2DM patients either with or without prior metformin monotherapy who initiate treatment with one of the 22 drug ingredients that comprise the DPP4 inhibitor, GLP1 receptor agonist, SGT2 inhibitor and sulfonylurea drug classes (Table 1). We do not consider thiazolidinediones given their known association with a risk of heart failure and bladder cancer [36, 37]. We describe specific definitions for exposure cohorts for each study in the following sections.

### 3.4 Exposure Comparators

#### 3.4.1 Class-vs-Class Study comparisons

The **Class-vs-Class** Study will construct four exposure cohorts for new-users of any drug ingredient within the four traditionally second-line drug classes in Table 1. Cohort entry (index date) for each patient is their first observed exposure to any drug ingredient for the four second-line drug classes. Consistent with an idealized target trial for T2DM therapy and cardiovascular risk [38, 39], inclusion criteria for patients based on the index date will include:

- T2DM diagnosis and no Type 1 or secondary diabetes mellitus diagnosis before the index date;
- At least 1 year of observation time before the index date (to improve new-user sensitivity); and
- No prior drug exposure to a comparator second-line or other antihyperglycemic agent (i.e. thiazolidinediones, acarbose, acetohexamide, bromocriptine, glibornuride, miglitol and nateglinide) or 30 days insulin exposure before index date.

We will construct and compare separately cohorts patients either with

- At least 3 months of metformin use before the index date,
- No prior metformin use before the index date.

In the first case, three months of metformin is consistent with ADA guidelines [40]. In the second case, we are interested in relative effectiveness and safety of these traditionally second-line agents in patients who initiate their treatments without first using metformin. We purposefully do not automatically exclude or restrict to patients with a history of myocardial infarction, stroke or other major cardiovascular events, which will allow us to report relative effectiveness and safety for individuals with both low or moderate and high cardiovascular risk. Likewise, we do not automatically exclude or restrict to individuals with severe renal impairment [41]. We will use cohort diagnostics, such as achieving covariate balance and clinical empirical equipoise between exposure cohorts (Section 4) and stakeholder input to guide the possible need to exclude other prior diagnoses, such as congestive heart failure, pancreatitis or cancer [41].

Appendix A.1 reports the complete OHDSI `ATLAS` cohort description for new-users of DDP4 inhibitors with prior metformin use. This description lists complete specification of cohort entry events, additional inclusion criteria, cohort exit events, and all associated standard OMOP CDM concept code sets used in the definition. We generate programmatically equivalent cohort definitions for new-others of each drug class with and without prior metformin use. `ATLAS` then automatically translates these definitions into network-deployable SQL source code. Appendix A.2 lists the inclusion criteria modifier for no prior metformin use.

Of note, the inclusion criteria do not directly incorporate quantitative measures of poor glycemic control, such as one or more elevated serum HbA1c measurements; such laboratory values are irregularly captured in large claims and even EHR data sources. Older ADA guidelines (but not since 2020 for patients with cardiovascular disease [42]) advise escalating to a second-line agent only when glycemic control is not met with metformin monotherapy, nicely mirroring our cohort design for our historical data. We will conduct sensitivity analyses involving available HbA1c measurements to demonstrate their bal-surements ≥ within 6 months before the index [39]. We will also conduct sensitivity

For each data source, we will then execute all 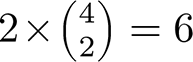 class comparisons forof patients strongly suggest data source-specific differences in prescribing practices that may introduce residual bias and sufficient samples sizes are required to construct effective propensity score models [43].

#### 3.4.2 Drug-vs-Drug Study comparisons

The **Drug-vs-Drug Study** will construct exposure cohorts for new-users of each drug ingredient in Table 1. We will apply the same cohort definition, inclusion criteria and patient count minimum as described in Section 3.4.1.

For each data source, we will then execute all 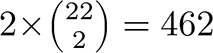 pairwise drug comparisons. While we will publicly report studies results for all pairwise comparisons, we will focus primary clinical interpretation and scientific publishing to the 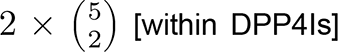 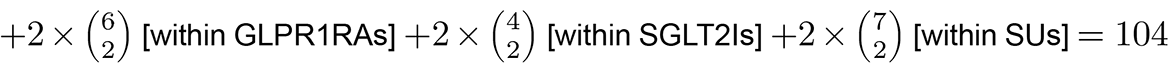 comparisons that pit drugs within the same class against each other, as well as across-class comparisons that stakeholders deem pertinent given their experiences.

Appendix A.5 reports the complete OHDSI `ATLAS` cohort description for new-users of aloglipitin with prior metformin use. Again, we programmatically construct all new-user drug-level cohort and automatically translate into SQL.

#### 3.4.3 Heterogeneity Study comparisons

The **Heterogeneity Study** will further stratify all 237 class- and drug-level exposure cohorts in Sections 3.4.1 and 3.4.2 by clinically important patient characteristics that modify cardiovascular risk or relative treatment heterogeneity to provide patient-focused treatment recommendations. These factors will include:

- Age (18 - 44 / 45 - 64 / ≥ 65 at the index date)
- Gender (women / men)
- Race (African American or black)
- Cardiovascular risk (low or moderate/high, defined by established cardiovascular disease at the index date)
- Renal impairment (at the index date)

We will define patients at high cardiovascular risk as those who fulfill at index date an established cardiovascular disease (CVD) definition that has been previously developed and validated for risk stratification among new-users of second-line T2DM agents [44]. Under this definition, established CVD means having at least 1 diagnosis code for a condition indicating cardiovascular disease, such as atherosclerotic vascular disease, cerebrovascular disease, ischemic heart disease or peripheral vascular disease, or having undergone at least 1 procedure indicating cardiovascular disease, such as percutaneous coronary intervention, coronary artery bypass graft or revascularization, any time on or prior to the exposure start. Likewise, we will define renal impairment through diagnosis codes for chronic kidney disease and end-stage renal disease, dialysis procedures, and laboratory measurements of estimated glomerular filtration rate, serum creatinine and urine albumin.

Appendix A.4 presents complete OHDSI `ATLAS` specifications for these subgroups, including all standard OMOP CDM concept codes defining cardiovascular risk and renal disease.

#### 3.4.4 Validation

We will validate exposure cohorts and aggregate drug utilization using comprehensive cohort characterization tools against both claims and EHR data sources. Chief among these tools stands OHDSI’s `CohortDiagnostic` package (github). For any cohort and data source mapped to OMOP CDM, this package systematically generates incidence new-user rates (stratified by age, gender, and calendar year), cohort characteristics (all comorbidities, drug use, procedures, health utilization) and the actual codes found in the data triggering the various rules in the cohort definitions. This can allow researchers and stakeholders to understand the heterogeneity of source coding for exposures and health outcomes as well as the impact of various inclusion criteria on overall cohort counts (details described in Section 4).

### 3.5 Outcomes

Across all data sources and pairwise exposure cohorts, we will assess relative risks of 32 cardiovascular and patient-centered outcomes (Table 3). Primary outcomes of interest are:

- 3-point major adverse cardiovascular events (MACE), including acute myocardial infarction, stroke, and sudden cardiac death, and
- 4-point MACE that additionally includes heart failure hospitalization.
- outcomes include:
- individual MACE components,
- acute renal failure,
- revascularization

In data sources with laboratory measurements, secondary outcomes further include:

- glycemic control, and
- measured renal dysfunction
- will also study second-line T2DM drug side-effects and safety concerns highlighted in the 2018 ADA guidelines [40] and from RCTs, including:
- abnormal weight change,
- genitourinary (GU) infection,
- various cancers, and
- hypoglycemia.

We will employ the same level of systematic rigor in studying outcomes regardless of their primary or secondary label.

A majority of outcome definitions have been previously implemented and validated in our own work [22,44–48] based heavily on prior development by others (see references in Table 3 [44–101]). To assess across-source consistency and general clinical validity, we will characterize outcome incidence, stratified by age, sex and index year for each data source.

### 3.6 Analysis

#### 3.6.1 Contemporary utilization of drug classes and individual agents

For all cohorts in the three studies, we will describe overall utilization as well as temporal trends in the use of each drug class and agents within the class. Further, we will eavaluate these trends in patient groups by age (18-44 / 45-64 / ≥ years), gender, race and For all cohorts in the three studies, we will describe overall utilization as well as temporal trends in the use of each drug class and agents within the class. Further, we will evaluate 65 geographic regions. Since the emergence of novel medications in the management of type 2 DM in 2014, there has been a rapid expansion in both the number of drug classes and individual agents. These data will provide insight into the current patterns of use and possible disparities. These data are critical to guide the real-world application of treatment decision pathways for the treatment of T2DM patients.

Specifically, we will calculate and validate aggregate drug utilization using the OHDSI’s CohortDiagnostic package against both claims and EHR data sources. The CohortDiagnostics package works in two steps: 1) Generate the utilization results and diagnostics against a data source and 2) Explore the generated utilization and diagnostics in a userfriendly graphical interface R-Shiny app. Through the interface, one can explore patient profiles of a random sample of subjects in a cohort. These diagnostics provide a consistent methodology to evaluate cohort definitions/phenotype algorithms across a variety of observational databases. This will enable researchers and stakeholders to become informed on the appropriateness of including specific data sources within analyses, exposing potential risks related to heterogeneity and variability in patient care delivery that, when not addressed in the design, could result in errors such as highly correlated covariates in propensity score matching of a target and a comparator cohort. Thus, the added value of this approach is two-fold in terms of exposing data quality for a study question and ensuring face validity checks are performed on proposed covariates to be used for balancing propensity scores.

#### 3.6.2 Relative risk of cardiovascular and patient-centered outcomes

For all three studies, we will execute a systematic process to estimate the relative risk of cardiovascular and patient-centered outcomes between new-users of second-line T2DM agents. The process will adjust for measured confounding, control from further residual (unmeasured) bias and accommodate important design choices to best emulate the nearly impossible to execute, idealized RCT that our stakeholders envision across data source populations, comparators, outcomes and subgroups.

To adjust for potential measured confounding and improve the balance between cohorts, we will build large-scale propensity score (PS) models [102] for each pairwise comparison and data source using a consistent data-driven process through regularized regression [31]. This process engineers a large set of predefined baseline patient characteristics, including age, gender, race, index month/year and other demographics and prior conditions, drug exposures, procedures, laboratory measurements and health service utilization behaviors, to provide the most accurate prediction of treatment and balance patient cohorts across many characteristics. Construction of condition, drug, procedures and observations include occurrences within 365, 180 and 30 days prior to index date and are aggregated at several SNOMED (conditions) and ingredient/ATC class (drugs) levels. Other demographic measures include comorbidity risk scores (Charlson, DCSI, CHADS2, CHAD2VASc). From prior work, feature counts have ranged in the 1,000s - 10,000s, and these large-scale PS models have outperformed hdPS [103] in simulation and real-world examples [31].

We will:

- Exclude patients who have experienced the outcome prior to their index date,
- Stratify and variable-ratio match patients by PS, and
- Use Cox proportional hazards models

to estimate hazard ratios (HRs) between alternative target and comparator treatments for the risk of each outcome in each data source. The regression will condition on the PS strata/matching-unit with treatment allocation as the sole explanatory variable and censor patients at the end of their time-at-risk (TAR) or data source observation period. We will prefer stratification over matching if both sufficiently balance patients (see Section 4), as the former optimizes patient inclusions and thus generalizability.

We will execute each comparison using three different TAR definitions, reflecting different and important causal contrasts:

- Intent-to-treat (TAR: index + 1 → end of observation) captures both direct treatment effects and (long-term) behavioral/treatment changes that initial assignment triggers [104];
- On-treatment-1 (TAR: index + 1 → treatment discontinuation) is more patient-centered [105] and captures direct treatment effect while allowing for escalation with additional T2DM agents; and
- On-treatment-2 (TAR: index + 1 → discontinuation or escalation with T2DM agents) carries the least possible confounding with other concurrent T2DM agents.

Our “on-treatment” is often called “per-protocol” [106]. Systematically executing with multiple causal contrasts enables us to identify potential biases that missing prescription data, treatment escalation and behavioral changes introduce, while preserving the ease of intent-to-treat interpretation and power if the data demonstrate them as unbiased. Appendix A.3 reports the modified cohort exit rule for the on-treatment-2 TAR.

We will aggregate HR estimates across non-overlapping data sources to produce metaanalytic estimates using a random-effects meta-analysis [107]. This classic meta-analysis assumes that per-data source likelihoods are approximately normally distributed [108]. This assumption fails when outcomes are rare as we expect for some safety events. Here, our recent research shows that as the number of data sources increases, the non-normality effect increases to where coverage of 95% confidence intervals (CIs) can be as low as 5%. To counter this, we will also apply a Bayesian meta-analysis model [109, 110] that neither assumes normality nor requires patient-level data sharing by building on composite likelihood methods [111] and enables us to introduce appropriate overlap weights between data sources.

Residual study bias from unmeasured and systematic sources often remains in observational studies even after controlling for measured confounding through PS-adjustment [32, 33]. For each comparison-outcome effect, we will conduct negative control (falsification) outcome experiments, where the null hypothesis of no effect is believed to be true, using approximately 100 controls. We identified these controls through a data-rich algorithm [112] that identifies prevalent OMOP condition concept occurrences that lack evidence of association with exposures in published literature, drug-product labeling and spontaneous reports, and were then adjudicated by clinical review. We previously validated 60 of the controls in LEGEND-HTN [22]. Appendix C lists these negative controls and their OMOP condition concept IDs.

Using the empirical null distributions from these experiments, we will calibrate each study effect HR estimate, its 95% CI and the *p*-value to reject the null hypothesis of no differential effect [34]. We will declare an HR as significantly different from no effect when its calibrated *p* < 0.05 without correcting for multiple testing. Finally, blinded to all trial results, study investigators will evaluate study diagnostics for all comparisons to assess if they were likely to yield unbiased estimates (Section 4).

#### 3.6.3 Sensitivity analyses and missingness

Because of the potential confounding effect of glycemic control at baseline between treatment choice and outcomes and to better understand the impact of limited glucose level measurements on effectiveness and safety estimation that arises in administrative claims and some EHR data, we will perform pre-specified sensitivity analyses for all studies within data sources that contain reliable glucose or hemoglobin A1c measurements. Within a study, for each exposure pair, we will first rebuild PS models where we additionally include baseline glucose or hemoglobin A1c measurements as patient characteristics, stratify or match patients under the new PS models that directly adjust for potential confounding by glycemic control and then estimate effectiveness and safety HRs.

A limitation of the Cox model is that no doubly robust procedure is believed to exist for estimating HRs, due to their non-collapsibility [113]. Doubly robust procedures combine baseline patient characteristic-adjusted outcome and PS models to control for confounding and, in theory, remain unbiased when either (but not necessarily both) model is correctly specified [114]. Doubly robust procedures do exist for hazard differences [113] and we will validate the appropriateness of our univariable Cox modeling by comparing estimate differences under an additive hazards model [116] with and without doubly robust-adjustment [117]. In practice, however, neither the outcome nor PS model is correctly specified, leading to systematic error in the observational setting.

Missing data of potential concern are patient demographics (gender, age, race) for our inclusion criteria. We will include only individuals whose baseline eligibility can be characterized that will most notably influence race subgroup assessments in the **Heterogeneity Study**. No further missing data can arise in our large-scale PS models because all features, with the exception of demographics, simply indicate the presence or absence of health records in a given time-period. Finally, we limit the impact of missing data, such as prescription information, relating to exposure time-at-risk by entertaining multiple definitions [29]. In all reports, we will clearly tabulate numbers of missing observations and patient attrition.

## 4 Sample Size and Study Power

Within each data source, we will execute all comparisons with ≥ 1,000 eligible patients per arm. Blinded to effect estimates, investigators and stakeholders will evaluate extensive study diagnostics for each comparison to assess reliability and generalizability, and only report risk estimates that pass [25, 35]. These diagnostics will include

1. Minimum detectable risk ratio (MDRR) as a typical proxy for power,
2. Preference score distributions to evaluate empirical equipoise10 and population generalizability,
3. Extensive patient characteristics to evaluate cohort balance before and after PS-adjustment,
4. Negative control calibration plots to assess residual bias, and
5. Kaplan-Meier plots to examine hazard ratio proportionality assumptions.

We will define cohorts to stand in empirical equipoise if the majority of patients carry preference scores between 0.3 and 0.7 and to achieve balance if all after-adjustment characteristics return absolute standardized mean differences 0.1 [118].

## 5 Strengths and Limitations

### 5.1 Strengths

LEGEND-T2DM is, to our knowledge, the largest and most comprehensive study to provide evidence about the comparative effectiveness and safety of second-line T2DM agents. The LEGEND-T2DM studies will encompass over 1 million patients initiating second-line T2DM agents across at least 13 databases from 5 countries and will examine all pairwise comparisons between the four second-line drug classes against a panel of TODO health outcomes. Through an international network, LEGEND-T2DM seeks to take advantage of disparate health databases drawn from different sources and across a range of countries and practice settings. These large-scale and unfiltered populations better represent real-world practice than the restricted study populations in prescribed treatment and follow-up settings from RCTs. Our use of the OMOP CDM allows extension of the LEGEND-T2DM experiment to future databases and allows replication of these results on licensable databases that were used in this experiment, while still maintaining patient privacy on patient-level data.

LEGEND-T2DM further advances the statistically rigorous and empirically validated methods we have developed in OHDSI that specifically address bias inherent in observational studies and allow for reliable causal inference. Patient characteristics and their treatment choices are likely to confound comparative effectiveness and safety estimates. Our approach combines active comparator new-user designs that emulate randomized clinical trials with large-scale propensity adjustment for measured confounding, a large set of negative control outcome experiments to address unmeasured and systematic bias, and full disclosure of hypotheses tested.

Each LEGEND-T2DM aim will represent evidence synthesis from a large number of bespoke studies across multiple data sources. Addressing questions one bespoke study at a time is prone to errors arising from multiple testing, random variation in effect estimates and publication bias. LEGEND-T2DM is designed to avoid these concerns through methodologic best practices [119] with full study diagnostics and external replication.

Through open science, LEGEND-T2DM will allow any interested investigators to engage as partners in our work at many levels. We will publicly develop all protocols and analytic code. This invites additional data custodians to participate in LEGEND-T2DM and enables others to modify and reuse our approach for other investigations. We will also host realtime access to all study result artifacts for outside analysis and interpretation. Such an open science framework ensures a feed-forward effect on other scientific contributions in the community. Collectively, LEGEND-T2DM will generate patient-centered, high quality, generalizable evidence that will transform the clinical management of T2DM through our active collaboration with patients, clinicians, and national medical societies. LEGENDT2DM will spur scientific innovation through the generation of open-source resources in data science.

### 5.2 Limitations

Even though many potential confounders will be included in these studies, there may be residual bias due to unmeasured or misspecified confounders, such as confounding by indication, differences in physician characteristics that may be associated with drug choice, concomitant use of other drugs started after the index date, and informative censoring at the end of the on-treatment periods. To minimize this risk, we will use methods to detect residual bias through a large number of negative and positive controls.

Ideal negative controls carry identical confounding between exposures and the outcome of interest [120]. The true confounding structure, however, is unknowable. Instead of attempting to find the elusive perfect negative control, we will rely on a large sample of controls that represent a wide range of confounding structures. If a study comparison proves to be unbiased for all negative controls, we can feel confident that it will also be unbiased for the outcome of interest. In our previous studies [22,25,121], using the active comparator, new-user cohort design we will employ here, we have observed minimal residual bias using negative controls. This stands in stark contrast to other designs such as the (nested) case-control that tends to show large residual bias because of incomparable exposure cohorts implied by the design [122].

Observed follow-up times are limited and variable, potentially reducing power to detect differences in effectiveness and safety and, further, misclassification of study variables is unavoidable in secondary use of health data, so it is possible to misclassify treatments, covariates, and outcomes. Based on our previous successful studies on antihypertensives, we do not expect differential misclassification, and therefore bias will most likely be towards the null. Finally, the electronic health record databases may be missing care episodes for patients due to care outside the respective health systems. Such bias, however, will also most likely be towards the null.

## 6 Protection of Human Subjects

LEGEND-T2DM does not involve human subjects research. The project does, however, use human data collected during routine healthcare provision. Most often the data are de-identified within data source. All data partners executing the LEGEND-T2DM studies within their data sources will have received institutional review board (IRB) approval or waiver for participation in accordance to their institutional governance prior to execution (see Table 4). LEGEND-T2DM executes across a federated and distributed data network, where analysis code is sent to participating data partners and only aggregate summary statistics are returned, with no sharing of patient-level data between organizations.

**Table 4:**
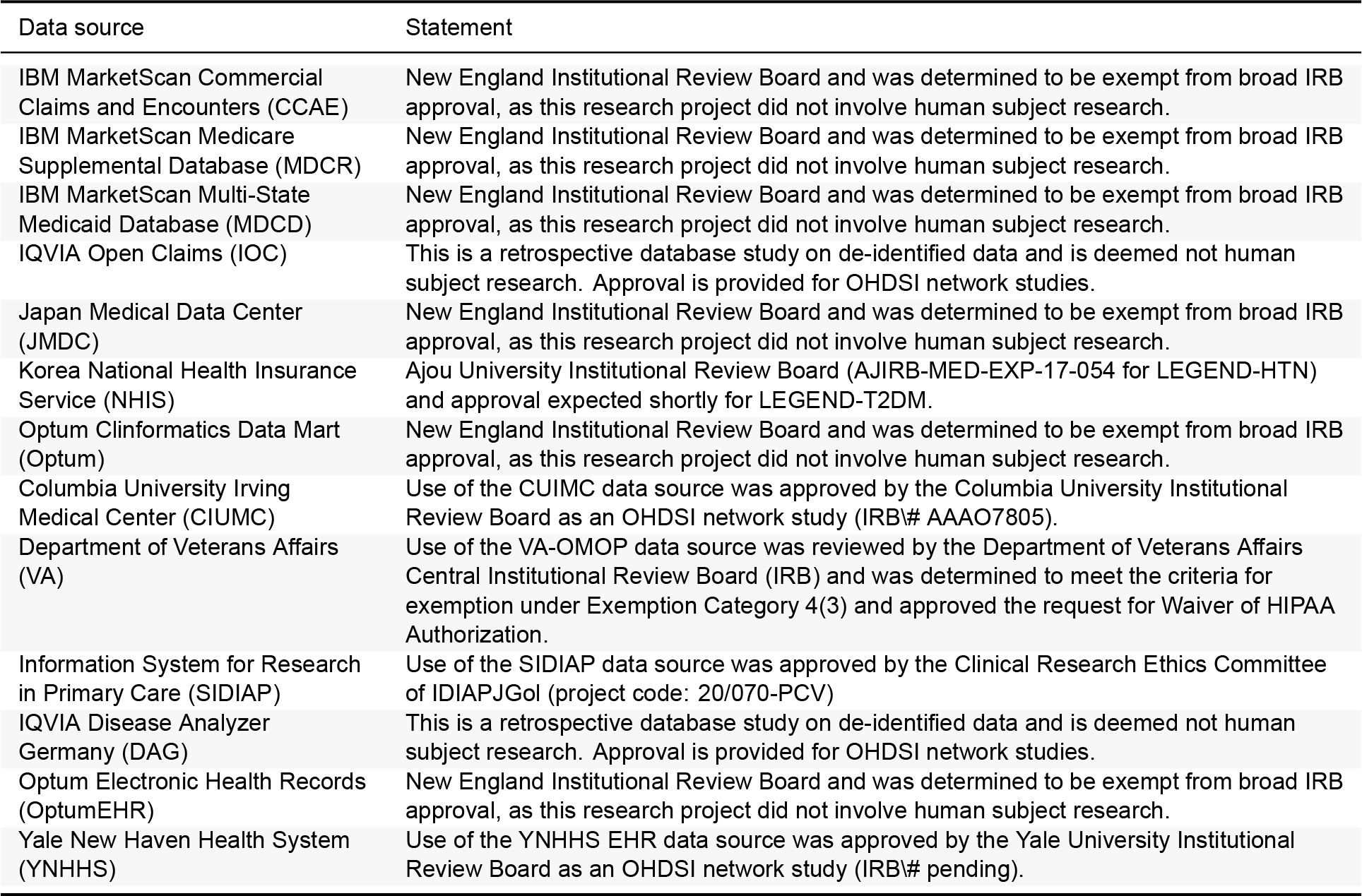
IRB approval or waiver statement from partners.

## 7 Management and Reporting of Adverse Events and Adverse Reactions

LEGEND-T2DM uses coded data that already exist in electronic databases. In these types of databases, it is not usually possible to link (i.e., identify a potential causal association between) a particular product and medical event for any specific individual. Thus, the minimum criteria for reporting an adverse event (i.e., identifiable patient, identifiable reporter, a suspect product and event) are not available and adverse events are not reportable as individual adverse event reports. The study results will be assessed for medically important findings.

## 8 Plans for Disseminating and Communicating Study Results

Open science aims to make scientific research, including its data process and software, and its dissemination, through publication and presentation, accessible to all levels of an inquiring society, amateur or professional [123] and is a governing principle of LEGENDT2DM. Open science delivers reproducible, transparent and reliable evidence. All aspects of LEGEND-T2DM (except private patient data) will be open and we will actively encourage other interested researchers, clinicians and patients to participate. This differs fundamentally from traditional studies that rarely open their analytic tools or share all result artifacts, and inform the community about hard-to-verify conclusions at completion.

### 8.1 Transparent and re-usable research tools

We will publicly register this protocol and announce its availability for feedback from stakeholders, the OHDSI community and within clinical professional societies. This protocol will link to open source code for all steps to generating diagnostics, effect estimates, figures and tables. Such transparency is possible because we will construct our studies on top of the OHDSI toolstack of open source software tools that are community developed and rigorously tested [25]. We will publicly host LEGEND-T2DM source code at (https://github.com/ohdsi-studies/LegendT2dm), allowing public contribution and review, and free re-use for anyone’s future research.

### 8.2 Continous sharing of results

LEGEND-T2DM embodies a new approach to generating evidence from healthcare data that overcome weaknesses in the current process of answering and publishing (or not) one question at a time. Generating evidence for thousands of research and control questions using a systematic process enables us to not only evaluate that process and the coherence and consistency of the evidence, but also to avoid *p*-hacking and publication bias [35]. We will store and openly communicate all of these results as they become available using a user-friendly web-based app that serves up all descriptive statistics, study diagnostics and effect estimates for each cohort comparison and outcome. Open access to this app will be through a general public facing LEGEND-T2DM webpage.

### 8.3 Scientific meetings and publications

We will deliver multiple presentations annually at scientific venues including the annual meetings of the American Diabetes Association, American College of Cardiology, American Heart Association and American Medical Informatics Association. We will also prepare multiple scientific publications for clinical, informatics and statistical journals.

### 8.4 General public

We believe in sharing our findings that will guide clinical care with the general public. LEGEND-T2DM will use social-media (Twitter) to facilitate this. With dedicated support from the OHDSI communications specialist, we will deliver regular press releases at key project stages, distributed via the extensive media networks of UCLA, Columbia and Yale.

## Data Availability

The study relies on a federated analytic model where the data reside with the source. The code for the study will be publicly available but the data can only be obtained in partnership with individuals and organizations included as data sources.

## Disclosures

This study is undertaken within Observational Health Data Sciences and Informatics (OHDSI), an open collaboration. **RK** is a founder of Evidence2Health, and receives grant funding from the US National Institutes of Health. **MJS** and **PBR** are employees of Janssen Research and Development and shareholders in John & Johnson. **GH** receives grant funding from the US National Institutes of Health and the US Food & Drug Administration and contracts from Janssen Research and Development. **HMK** receives grants from the US Food & Drug Administration, Medtronics and Janssen Research and Development, is co-founder of HugoHealth and chairs the Cardiac Scientific Advisory Board for UnitedHealth. **MAS** receives grant funding from the US National Institutes of Health, the US Department of Veterans Affairs and the US Food & Drug Administration and contracts from Janssen Research and Development and IQVIA.

## List of Abbreviations

CDM: Common data model
DPP4: Dipeptidyl peptidase-4
GLP1: Glucagon-like peptide-1
IRB: Institutional review board
LEGEND: Large-scale Evidence Generation and Evaluation across a Network of Databases
MACE: Major adverse cardiovascular event
MDRR: Minimum detectable risk ratio
OHDSI: Observational Health Data Science and Informatics
OMOP: Observational Medical Outcomes Partnership
PS: Propensity score
RCT: Randomized controlled trial
SGLT2: Sodium-glucose co-transporter-2
T2DM: Type 2 diabetes mellitus

## Appendix

### A Exposure Cohort Definitions

#### A.1 Class-vs-Class Exposure (DPP4 New-User) Cohort / OT1

##### A.1.1 Cohort Entry Events

People with continuous observation of 365 days before event may enter the cohort when observing any of the following:

1. drug exposure of ‘DPP4 inhibitors’ for the first time in the person’s history.

Limit cohort entry events to the earliest event per person. Restrict entry events to with all of the following criteria:

1. with the following event criteria: who are >= 18 years old.
2. having at least 1 condition occurrence of ‘Type 2 diabetes mellitus’, starting anytime on or before cohort entry start date; allow events outside observation period.
3. having no condition occurrences of ‘Type 1 diabetes mellitus’, starting anytime on or before cohort entry start date; allow events outside observation period.
4. having no condition occurrences of ‘Secondary diabetes mellitus’, starting anytime on or before cohort entry start date; allow events outside observation period.

##### A.1.2 Additional Inclusion Criteria

- No prior GLP-1 receptor agonist exposure

Entry events having no drug exposures of ‘GLP-1 receptor agonists’, starting anytime on or before cohort entry start date; allow events outside observation period.

- No prior SGLT-2 inhibitor exposure

Entry events having no drug exposures of ‘SGLT2 inhibitors’, starting anytime on or before cohort entry start date; allow events outside observation period.

- No prior SU exposure

Entry events having no drug exposures of ‘Sulfonylureas’, starting anytime on or before cohort entry start date; allow events outside observation period.

- No prior other anti-diabetic exposure

Entry events having no drug exposures of ‘Other anti-diabetics’, starting anytime on or before cohort entry start date; allow events outside observation period.

- Prior metformin use

Entry events with any of the following criteria:

1. having at least 1 drug era of ‘Metformin’, starting anytime up to 90 days before cohort entry start date; allow events outside observation period; with era length >= 90 days.
2. having at least 3 drug exposures of ‘Metformin’, starting anytime on or before cohort entry start date; allow events outside observation period.
3. No prior insulin use or combo initiation: Proxy for < 30 days drug era anytime before index and no combination use on index

Entry events with all of the following criteria:

1. having no drug eras of ‘Insulin’, starting anytime up to 30 days before cohort entry start date; allow events outside observation period; with era length > 30 days.
2. having no drug eras of ‘Insulin’, starting between 30 days before and 0 days after cohort entry start date; allow events outside observation period.

##### A.1.3 Cohort Exit

The cohort end date will be based on a continuous exposure to ‘DPP4 inhibitors’: allowing 30 days between exposures, adding 0 days after exposure ends, and using days supply and exposure end date for exposure duration.

##### A.1.4 Cohort Eras

Entry events will be combined into cohort eras if they are within 0 days of each other.

##### A.1.5 Concept: DPP4 inhibitors

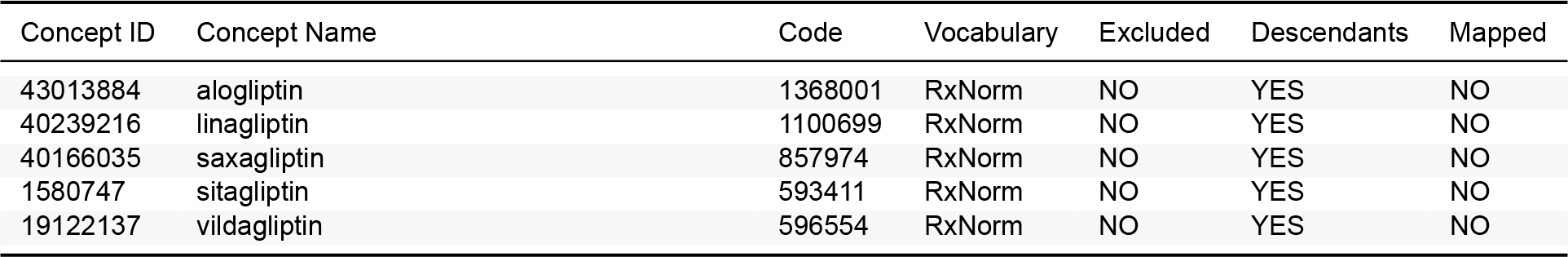

| Concept ID | Concept Name | Code | Vocabulary | Excluded | Descendants | Mapped |
| --- | --- | --- | --- | --- | --- | --- |
| 43013884 | alogliptin | 1368001 | RxNorm | NO | YES | NO |
| 40239216 | linagliptin | 1100699 | RxNorm | NO | YES | NO |
| 40166035 | saxagliptin | 857974 | RxNorm | NO | YES | NO |
| 1580747 | sitagliptin | 593411 | RxNorm | NO | YES | NO |
| 19122137 | vildagliptin | 596554 | RxNorm | NO | YES | NO |

##### A.1.6 Concept: GLP-1 receptor agonists

| Concept ID | Concept Name | Code | Vocabulary | Excluded | Descendants | Mapped |
| --- | --- | --- | --- | --- | --- | --- |
| 44816332 | albiglutide | 1534763 | RxNorm | NO | YES | NO |
| 45774435 | dulaglutide | 1551291 | RxNorm | NO | YES | NO |
| 1583722 | exenatide | 60548 | RxNorm | NO | YES | NO |
| 40170911 | liraglutide | 475968 | RxNorm | NO | YES | NO |
| 44506754 | lixisenatide | 1440051 | RxNorm | NO | YES | NO |
| 793143 | semaglutide | 1991302 | RxNorm | NO | YES | NO |

##### A.1.7 Concept: SGLT2 inhibitors

| Concept ID | Concept Name | Code | Vocabulary | Excluded | Descendants | Mapped |
| --- | --- | --- | --- | --- | --- | --- |
| 43526465 | canagliflozin | 1373458 | RxNorm | NO | YES | NO |
| 44785829 | dapagliflozin | 1488564 | RxNorm | NO | YES | NO |
| 45774751 | empagliflozin | 1545653 | RxNorm | NO | YES | NO |
| 793293 | ertugliflozin | 1992672 | RxNorm | NO | YES | NO |

##### A.1.8 Concept: Sulfonylureas

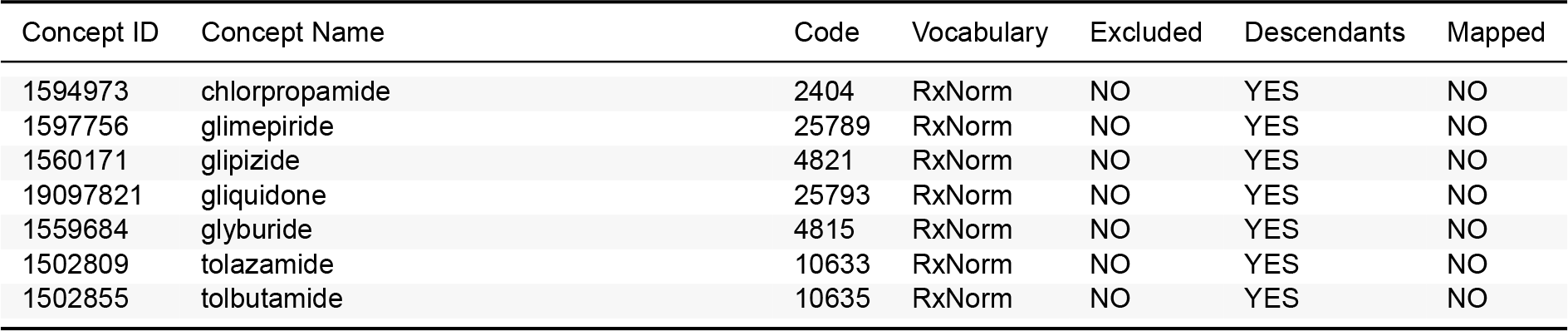

| Concept ID | Concept Name | Code | Vocabulary | Excluded | Descendants | Mapped |
| --- | --- | --- | --- | --- | --- | --- |
| 1594973 | chlorpropamide | 2404 | RxNorm | NO | YES | NO |
| 1597756 | glimepiride | 25789 | RxNorm | NO | YES | NO |
| 1560171 | glipizide | 4821 | RxNorm | NO | YES | NO |
| 19097821 | gliquidone | 25793 | RxNorm | NO | YES | NO |
| 1559684 | glyburide | 4815 | RxNorm | NO | YES | NO |
| 1502809 | tolazamide | 10633 | RxNorm | NO | YES | NO |
| 1502855 | tolbutamide | 10635 | RxNorm | NO | YES | NO |

##### A.1.9 Concept: Other anti-diabetics

| Concept ID | Concept Name | Code | Vocabulary | Excluded | Descendants | Mapped |
| --- | --- | --- | --- | --- | --- | --- |
| 1529331 | acarbose | 16681 | RxNorm | NO | YES | NO |
| 1530014 | acetohexamide | 173 | RxNorm | NO | YES | NO |
| 730548 | bromocriptine | 1760 | RxNorm | NO | YES | NO |
| 19033498 | carbutamide | 2068 | RxNorm | NO | YES | NO |
| 19001409 | glibornuride | 102846 | RxNorm | NO | YES | NO |
| 19059796 | gliclazide | 4816 | RxNorm | NO | YES | NO |
| 19001441 | glymidine | 102848 | RxNorm | NO | YES | NO |
| 1510202 | miglitol | 30009 | RxNorm | NO | YES | NO |
| 1502826 | nateglinide | 274332 | RxNorm | NO | YES | NO |
| 1525215 | pioglitazone | 33738 | RxNorm | NO | YES | NO |
| 1516766 | repaglinide | 73044 | RxNorm | NO | YES | NO |
| 1547504 | rosiglitazone | 84108 | RxNorm | NO | YES | NO |
| 1515249 | troglitazone | 72610 | RxNorm | NO | YES | NO |

##### A.1.10 Concept: Insulin

| Concept ID | Concept Name | Code | Vocabulary | Excluded | Descendants | Mapped |
| --- | --- | --- | --- | --- | --- | --- |
| 1596977 | insulin, regular, human | 253182 | RxNorm | NO | YES | NO |
| 1550023 | insulin lispro | 86009 | RxNorm | NO | YES | NO |
| 1567198 | insulin aspart, human | 51428 | RxNorm | NO | YES | NO |
| 1502905 | insulin glargine | 274783 | RxNorm | NO | YES | NO |
| 1513876 | insulin lispro protamine, human | 314684 | RxNorm | NO | YES | NO |
| 1531601 | insulin aspart protamine, human | 352385 | RxNorm | NO | YES | NO |
| 1586346 | insulin, regular, pork | 221109 | RxNorm | NO | YES | NO |
| 1544838 | insulin glulisine, human | 400008 | RxNorm | NO | YES | NO |
| 1516976 | insulin detemir | 139825 | RxNorm | NO | YES | NO |
| 1590165 | insulin, regular, beef-pork | 235275 | RxNorm | NO | YES | NO |
| 1513849 | lente insulin, human | 314683 | RxNorm | NO | YES | NO |
| 1562586 | lente insulin, pork | 93108 | RxNorm | NO | YES | NO |
| 1588986 | insulin human, rDNA origin | 631657 | RxNorm | NO | YES | NO |
| 1513843 | lente insulin, beef-pork | 314682 | RxNorm | NO | YES | NO |
| 1586369 | ultralente insulin, human | 221110 | RxNorm | NO | YES | NO |
| 35605670 | insulin argine | 1740938 | RxNorm | NO | YES | NO |
| 35602717 | insulin degludec | 1670007 | RxNorm | NO | YES | NO |
| 21600713 | INSULINS AND ANALOGUES | A10A | ATC | NO | YES | NO |
| 19078608 | insulin, protamine zinc, beef-pork 100 UNT/ML Injectable Suspension | 311053 | RxNorm | NO | YES | NO |

##### A.1.11 Concept: Metformin

| Concept ID | Concept Name | Code | Vocabulary | Excluded | Descendants | Mapped |
| --- | --- | --- | --- | --- | --- | --- |
| 1503297 | metformin | 6809 | RxNorm | NO | YES | NO |

##### A.1.12 Concept: Secondary diabetes mellitus

| Concept ID | Concept Name | Code | Vocabulary | Excluded | Descendants | Mapped |
| --- | --- | --- | --- | --- | --- | --- |
| 195771 | Secondary diabetes mellitus | 8801005 | SNOMED | NO | YES | NO |

##### A.1.13 Concept: Type 1 diabetes mellitus

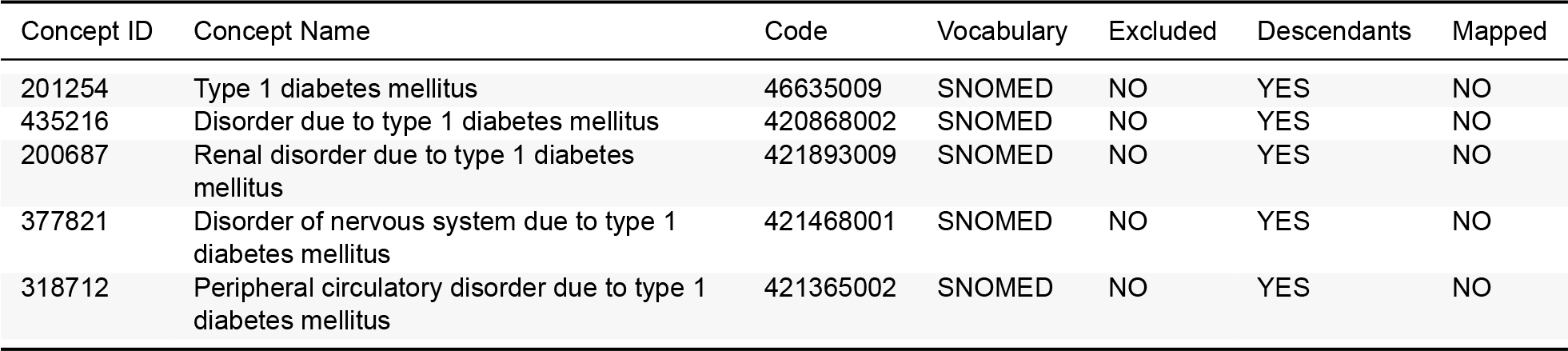

| Concept ID | Concept Name | Code | Vocabulary | Excluded | Descendants | Mapped |
| --- | --- | --- | --- | --- | --- | --- |
| 201254 | Type 1 diabetes mellitus | 46635009 | SNOMED | NO | YES | NO |
| 435216 | Disorder due to type 1 diabetes mellitus | 420868002 | SNOMED | NO | YES | NO |
| 200687 | Renal disorder due to type 1 diabetes mellitus | 421893009 | SNOMED | NO | YES | NO |
| 377821 | Disorder of nervous system due to type 1 diabetes mellitus | 421468001 | SNOMED | NO | YES | NO |
| 318712 | Peripheral circulatory disorder due to type 1 diabetes mellitus | 421365002 | SNOMED | NO | YES | NO |

##### A.1.14 Concept: Type 2 diabetes mellitus

| Concept ID | Concept Name | Code | Vocabulary | Excluded | Descendants | Mapped |
| --- | --- | --- | --- | --- | --- | --- |
| 201826 | Type 2 diabetes mellitus | 44054006 | SNOMED | NO | YES | NO |
| 443734 | Ketoacidosis due to type 2 diabetes mellitus | 421750000 | SNOMED | NO | YES | NO |
| 443767 | Disorder of eye due to diabetes mellitus | 25093002 | SNOMED | NO | YES | NO |
| 192279 | Disorder of kidney due to diabetes mellitus | 127013003 | SNOMED | NO | YES | NO |
| 443735 | Coma due to diabetes mellitus | 420662003 | SNOMED | NO | YES | NO |
| 376065 | Disorder of nervous system due to type 2 diabetes mellitus | 421326000 | SNOMED | NO | YES | NO |
| 443729 | Peripheral circulatory disorder due to type 2 diabetes mellitus | 422166005 | SNOMED | NO | YES | NO |
| 443732 | Disorder due to type 2 diabetes mellitus | 422014003 | SNOMED | NO | YES | NO |

#### A.2 Metformin Use Modifier

##### A.2.1 No prior metformin use

Entry events having no drug eras of ‘Metformin’, starting anytime on or before cohort entry start date; allow events outside observation period.

#### A.3 Escalation Exit Criteria

The cohort end date will be based on a continuous exposure to ‘DPP4 inhibitors’: allowing 30 days between exposures, adding 0 days after exposure ends, and using days supply and exposure end date for exposure duration.

The person also exists the cohort when encountering any of the following events:

1. drug exposures of ‘All alternative target exposures’.
2. drug exposures of ‘Other anti-diabetics’.
3. drug eras of ‘Insulin’, with era length > 30 days.

##### A.3.1 Concept: All alternative target exposures

| Concept ID | Concept Name | Code | Vocabulary | Excluded | Descendants | Mapped |
| --- | --- | --- | --- | --- | --- | --- |
| 44816332 | albiglutide | 1534763 | RxNorm | NO | YES | NO |
| 43526465 | canagliflozin | 1373458 | RxNorm | NO | YES | NO |
| 1594973 | chlorpropamide | 2404 | RxNorm | NO | YES | NO |
| 44785829 | dapagliflozin | 1488564 | RxNorm | NO | YES | NO |
| 45774435 | dulaglutide | 1551291 | RxNorm | NO | YES | NO |
| 45774751 | empagliflozin | 1545653 | RxNorm | NO | YES | NO |
| 793293 | ertugliflozin | 1992672 | RxNorm | NO | YES | NO |
| 1583722 | exenatide | 60548 | RxNorm | NO | YES | NO |
| 1597756 | glimepiride | 25789 | RxNorm | NO | YES | NO |
| 1560171 | glipizide | 4821 | RxNorm | NO | YES | NO |
| 19097821 | gliquidone | 25793 | RxNorm | NO | YES | NO |
| 1559684 | glyburide | 4815 | RxNorm | NO | YES | NO |
| 40170911 | liraglutide | 475968 | RxNorm | NO | YES | NO |
| 44506754 | lixisenatide | 1440051 | RxNorm | NO | YES | NO |
| 793143 | semaglutide | 1991302 | RxNorm | NO | YES | NO |
| 1502809 | tolazamide | 10633 | RxNorm | NO | YES | NO |
| 1502855 | tolbutamide | 10635 | RxNorm | NO | YES | NO |

#### A.4 Heterogenity Study Inclusion Criteria

##### A.4.1 Lower age group

Entry events with the following event criteria: who are < 45 years old.

##### A.4.2 Middle age group

Entry events with all of the following criteria:

1. with the following event criteria: who are >= 45 years old.
2. with the following event criteria: who are < 65 years old.

##### A.4.3 Older age group

Entry events with the following event criteria: who are >= 65 years old.

##### A.4.4 Female stratum

Entry events with the following event criteria: who are female.

##### A.4.5 Male stratum

Entry events with the following event criteria: who are male.

##### A.4.6 Race stratum

Entry events with the following event criteria: race is: “black or african american”, “black”, “african american”, “african”, “bahamian”, “barbadian”, “dominican”, “dominica islander”, “haitian”, “jamaican”, “tobagoan”, “trinidadian” or “west indian”.

##### A.4.7 Low cardiovascular risk

Entry events with all of the following criteria:

1. having no condition occurrences of ‘Conditions indicating established cardiovascular disease’, starting anytime on or before cohort entry start date; allow events outside observation period.
2. having no procedure occurrences of ‘Procedures indicating established cardiovascular disease’, starting anytime on or before cohort entry start date; allow events outside observation period.

##### A.4.8 Higher cardiovascular risk

Entry events with any of the following criteria:

1. having at least 1 condition occurrence of ‘Conditions indicating established cardiovascular disease’, starting anytime on or before cohort entry start date; allow events outside observation period.
2. having at least 1 procedure occurrence of ‘Procedures indicating established cardiovascular disease’, starting anytime on or before cohort entry start date; allow events outside observation period.

##### A.4.9 Concept: Conditions indicating established cardiovascular disease

| Concept ID | Concept Name | Code | Vocabulary | Excluded | Descendants | Mapped |
| --- | --- | --- | --- | --- | --- | --- |
| 319844 | Acute ischemic heart disease | 413439005 | SNOMED | NO | YES | NO |
| 321318 | Angina pectoris | 194828000 | SNOMED | NO | YES | NO |
| 4124841 | Aortic bifurcation syndrome | 233972005 | SNOMED | YES | YES | NO |
| 312337 | Arterial embolus and thrombosis | 266262004 | SNOMED | NO | YES | NO |
| 4278217 | Arterial thrombosis | 65198009 | SNOMED | NO | YES | NO |
| 40484167 | Arteriosclerosis of artery of extremity | 443971004 | SNOMED | NO | YES | NO |
| 318443 | Arteriosclerotic vascular disease | 72092001 | SNOMED | NO | YES | NO |
| 314659 | Arteritis | 52089001 | SNOMED | NO | NO | NO |
| 40479625 | Atherosclerosis of artery | 441574008 | SNOMED | NO | YES | NO |
| 40484541 | Atherosclerosis of autologous vein bypass graft of limb | 442693003 | SNOMED | YES | YES | NO |
| 312902 | Benign intracranial hypertension | 68267002 | SNOMED | YES | YES | NO |
| 4288310 | Carotid artery obstruction | 69798007 | SNOMED | YES | YES | NO |
| 372924 | Cerebral artery occlusion | 20059004 | SNOMED | NO | YES | NO |
| 376713 | Cerebral hemorrhage | 274100004 | SNOMED | NO | YES | NO |
| 381591 | Cerebrovascular disease | 62914000 | SNOMED | NO | YES | NO |
| 316494 | Cerebrovascular disorder in the puerperium | 6594005 | SNOMED | YES | YES | NO |
| 315286 | Chronic ischemic heart disease | 413838009 | SNOMED | NO | YES | NO |
| 44782819 | Chronic occlusion of artery of extremity | 698816006 | SNOMED | NO | YES | NO |
| 4313767 | Chronic peripheral venous hypertension | 423674003 | SNOMED | YES | YES | NO |
| 372721 | Congenital anomaly of cerebrovascular system | 65587001 | SNOMED | YES | YES | NO |
| 316995 | Coronary occlusion | 63739005 | SNOMED | NO | YES | NO |
| 134057 | Disorder of cardiovascular system | 49601007 | SNOMED | NO | NO | NO |
| 40480453 | Disorder of vein of lower extremity | 441739009 | SNOMED | YES | YES | NO |
| 46272492 | Dissection of artery | 710864009 | SNOMED | YES | YES | NO |
| 4324690 | Fracture of skull | 71642004 | SNOMED | YES | YES | NO |
| 441246 | Hemangioma of intracranial structure | 93468003 | SNOMED | YES | YES | NO |
| 380113 | Hemorrhage in optic nerve sheaths | 14460007 | SNOMED | YES | YES | NO |
| 192763 | Injury of blood vessel | 57662003 | SNOMED | YES | YES | NO |
| 4275428 | Injury of vein | 64583005 | SNOMED | YES | YES | NO |
| 442774 | Intermittent claudication | 63491006 | SNOMED | NO | YES | NO |
| 439847 | Intracranial hemorrhage | 1386000 | SNOMED | NO | YES | NO |
| 434056 | Late effects of cerebrovascular disease | 195239002 | SNOMED | NO | YES | NO |
| 4146311 | Leriche's syndrome | 307816004 | SNOMED | NO | YES | NO |
| 4329847 | Myocardial infarction | 22298006 | SNOMED | NO | YES | NO |
| 4296029 | Periarteritis | 76805007 | SNOMED | NO | YES | NO |
| 260841 | Perinatal subarachnoid hemorrhage | 21202004 | SNOMED | YES | YES | NO |
| 317309 | Peripheral arterial occlusive disease | 399957001 | SNOMED | NO | YES | NO |
| 321822 | Peripheral vascular disorder due to diabetes mellitus | 421895002 | SNOMED | NO | YES | NO |
| 313928 | Peripheral vascular complication | 10596002 | SNOMED | NO | YES | NO |
| 321052 | Peripheral vascular disease | 400047006 | SNOMED | NO | NO | NO |
| 44782775 | Peripheral vascular disease associated with another disorder | 34881000119105 | SNOMED | NO | YES | NO |
| 318137 | Phlebitis and thrombophlebitis of intracranial sinuses | 192753009 | SNOMED | YES | YES | NO |
| 441039 | Phlebitis of lower limb vein | 312588002 | SNOMED | NO | YES | NO |
| 4067424 | Polyarteritis | 20258000 | SNOMED | NO | YES | NO |
| 320749 | Polyarteritis nodosa | 155441006 | SNOMED | YES | YES | NO |
| 443239 | Precerebral arterial occlusion | 266253001 | SNOMED | NO | YES | NO |
| 440417 | Pulmonary embolism | 59282003 | SNOMED | YES | YES | NO |
| 4318842 | Renal vasculitis | 95578000 | SNOMED | NO | YES | NO |
| 380943 | Rupture of syphilitic cerebral aneurysm | 186893003 | SNOMED | YES | YES | NO |
| 432923 | Subarachnoid hemorrhage | 21454007 | SNOMED | NO | YES | NO |
| 439040 | Subdural hemorrhage | 35486000 | SNOMED | NO | YES | NO |
| 320741 | Thrombophlebitis | 64156001 | SNOMED | YES | YES | NO |
| 4141106 | Thrombosis of arteries of the extremities | 33591000 | SNOMED | NO | YES | NO |
| 4132546 | Traumatic brain injury | 127295002 | SNOMED | YES | YES | NO |
| 4194610 | Trunk arterial embolus | 312593004 | SNOMED | NO | YES | NO |
| 318169 | Varicose veins of lower extremity | 72866009 | SNOMED | YES | YES | NO |
| 4189293 | Vascular disorder of lower extremity | 373408007 | SNOMED | NO | YES | NO |
| 443752 | Ventricular hemorrhage | 23276006 | SNOMED | YES | YES | NO |
| 432346 | Dissection of vertebral artery | 230730001 | SNOMED | YES | YES | NO |

##### A.4.10 Concept: Procedures indicating established cardiovascular disease

| Concept ID | Concept Name | Code | Vocabulary | Excluded | Descendants | Mapped |
| --- | --- | --- | --- | --- | --- | --- |
| 4150819 | Operative procedure on coronary artery | 31413008 | SNOMED | NO | YES | NO |
| 4331725 | Operative procedure on artery of extremity | 22701007 | SNOMED | NO | YES | NO |

##### A.4.11 Without renal impairment

Entry events having no condition occurrences of ‘Renal impairment’, starting anytime on or before cohort entry start date; allow events outside observation period.

##### A.4.12 Renal impairment

Entry events having at least 1 condition occurrence of ‘Renal impairment’, starting anytime on or before cohort entry start date; allow events outside observation period.

##### A.4.13 Concept: Renal impairment

| Concept ID | Concept Name | Code | Vocabulary | Excluded | Descendants | Mapped |
| --- | --- | --- | --- | --- | --- | --- |
| 4030518 | Renal impairment | 236423003 | SNOMED | NO | YES | NO |

#### A.5 Drug-vs-Drug Exposure (Alogliptin New-User) Cohort / OT1

##### A.5.1 Cohort Entry Events

People with continuous observation of 365 days before event may enter the cohort when observing any of the following:

1. drug exposure of ‘alogliptin’ for the first time in the person’s history.

Limit cohort entry events to the earliest event per person. Restrict entry events to with all of the following criteria:

1. with the following event criteria: who are >= 18 years old.
2. having at least 1 condition occurrence of ‘Type 2 diabetes mellitus’, starting anytime on or before cohort entry start date; allow events outside observation period.
3. having no condition occurrences of ‘Type 1 diabetes mellitus’, starting anytime on or before cohort entry start date; allow events outside observation period.
4. having no condition occurrences of ‘Secondary diabetes mellitus’, starting anytime on or before cohort entry start date; allow events outside observation period.

##### A.5.2 Additional Inclusion Criteria

- No prior with-in class exposure

Entry events having no drug exposures of ‘DPP4 inhibitors excluding alogliptin’, starting anytime on or before cohort entry start date; allow events outside observation period.

- No prior GLP-1 receptor agonist exposure

Entry events having no drug exposures of ‘GLP-1 receptor agonists’, starting anytime on or before cohort entry start date; allow events outside observation period.

- No prior SGLT-2 inhibitor exposure

Entry events having no drug exposures of ‘SGLT2 inhibitors’, starting anytime on or before cohort entry start date; allow events outside observation period.

- No prior SU exposure

Entry events having no drug exposures of ‘Sulfonylureas’, starting anytime on or before cohort entry start date; allow events outside observation period.

- No prior other anti-diabetic exposure

Entry events having no drug exposures of ‘Other anti-diabetics’, starting anytime on or before cohort entry start date; allow events outside observation period.

- Prior metformin use

Entry events with any of the following criteria:

1. having at least 1 drug era of ‘Metformin’, starting anytime up to 90 days before cohort entry start date; allow events outside observation period; with era length >= 90 days.
2. having at least 3 drug exposures of ‘Metformin’, starting anytime on or before cohort entry start date; allow events outside observation period.
3. No prior insulin use or combo initiation: Proxy for < 30 days drug era anytime before index and no combination use on index

Entry events having no drug eras of ‘Insulin’, starting anytime on or before cohort entry start date; allow events outside observation period; with era length > 30 days.

##### A.5.3 Cohort Exit

The cohort end date will be based on a continuous exposure to ‘alogliptin’: allowing 30 days between exposures, adding 0 days after exposure ends, and using days supply and exposure end date for exposure duration.

##### A.5.4 Cohort Eras

Entry events will be combined into cohort eras if they are within 0 days of each other.

##### A.5.5 Concept: alogliptin

| Concept ID | Concept Name | Code | Vocabulary | Excluded | Descendants | Mapped |
| --- | --- | --- | --- | --- | --- | --- |
| 43013884 | alogliptin | 1368001 | RxNorm | NO | YES | NO |

##### A.5.6 Concept: DPP4 inhibitors excluding alogliptin

| Concept ID | Concept Name | Code | Vocabulary | Excluded | Descendants | Mapped |
| --- | --- | --- | --- | --- | --- | --- |
| 40239216 | linagliptin | 1100699 | RxNorm | NO | YES | NO |
| 40166035 | saxagliptin | 857974 | RxNorm | NO | YES | NO |
| 1580747 | sitagliptin | 593411 | RxNorm | NO | YES | NO |
| 19122137 | vildagliptin | 596554 | RxNorm | NO | YES | NO |

### B Outcome Cohort Definitions

#### B.1 3-point MACE

##### B.1.1 Cohort Entry Events

People may enter the cohort when observing any of the following:

1. condition occurrences of ‘Acute myocardial Infarction’.
2. condition occurrences of ‘Sudden cardiac death’.
3. condition occurrences of ‘Ischemic stroke’.
4. condition occurrences of ‘Intracranial bleed Hemorrhagic stroke’.

Restrict entry events to having at least 1 visit occurrence of ‘Inpatient or ER visit’, starting anytime on or before cohort entry start date and ending between 0 days before and all days after cohort entry start date.

##### B.1.2 Cohort Exit

The cohort end date will be offset from index event’s start date plus 7 days.

#### B.1.3 Cohort Eras

Entry events will be combined into cohort eras if they are within 180 days of each other.

##### B.1.4 Concept: Inpatient or ER visit

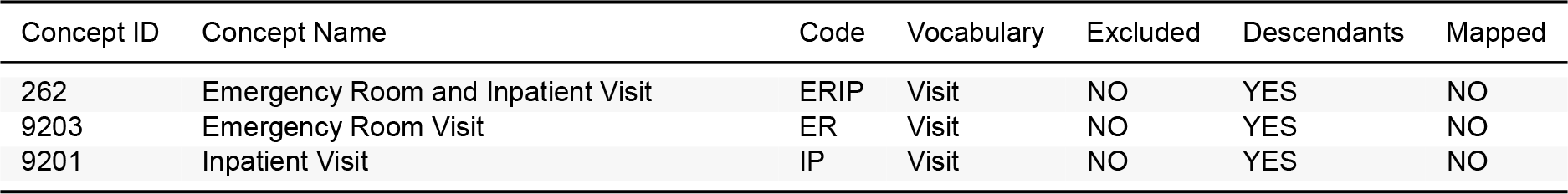

##### B.1.5 Concept: Acute myocardial Infarction

| Concept ID | Concept Name | Code | Vocabulary | Excluded | Descendants | Mapped |
| --- | --- | --- | --- | --- | --- | --- |
| 4329847 | Myocardial infarction | 22298006 | SNOMED | NO | YES | NO |
| 314666 | Old myocardial infarction | 1755008 | SNOMED | YES | YES | NO |

##### B.1.6 Concept: Sudden cardiac death

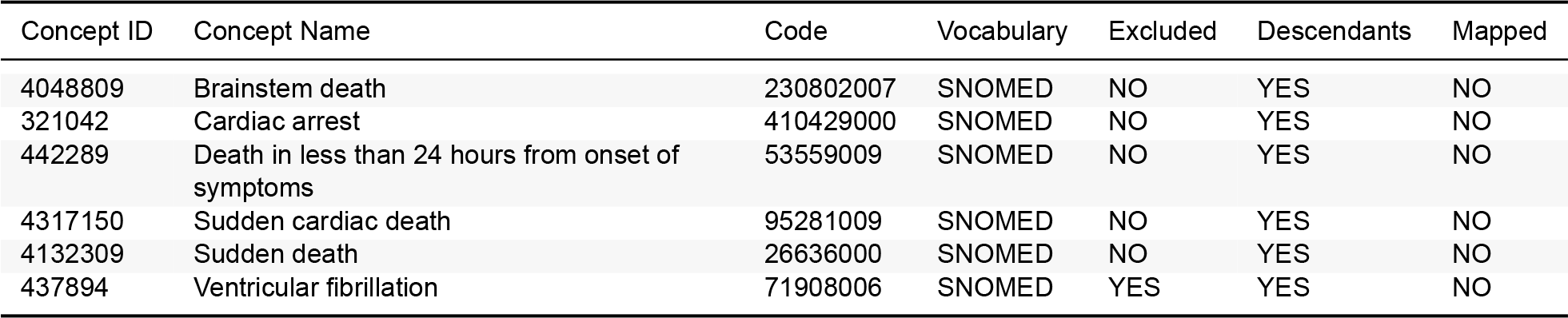

##### B.1.7 Concept: Ischemic stroke

| Concept ID | Concept Name | Code | Vocabulary | Excluded | Descendants | Mapped |
| --- | --- | --- | --- | --- | --- | --- |
| 372924 | Cerebral artery occlusion | 20059004 | SNOMED | NO | NO | NO |
| 375557 | Cerebral embolism | 75543006 | SNOMED | NO | NO | NO |
| 443454 | Cerebral infarction | 432504007 | SNOMED | NO | YES | NO |
| 441874 | Cerebral thrombosis | 71444005 | SNOMED | NO | NO | NO |

##### B.1.8 Concept: Intracranial bleed Hemorrhagic stroke

| Concept ID | Concept Name | Code | Vocabulary | Excluded | Descendants | Mapped |
| --- | --- | --- | --- | --- | --- | --- |
| 376713 | Cerebral hemorrhage | 274100004 | SNOMED | NO | NO | NO |
| 439847 | Intracranial hemorrhage | 1386000 | SNOMED | NO | NO | NO |
| 432923 | Subarachnoid hemorrhage | 21454007 | SNOMED | NO | NO | NO |
| 43530727 | Spontaneous cerebral hemorrhage | 291571000119106 | SNOMED | NO | NO | NO |
| 4148906 | Spontaneous subarachnoid hemorrhage | 270907008 | SNOMED | NO | NO | NO |

##### B.1.9 Concept: Heart Failure

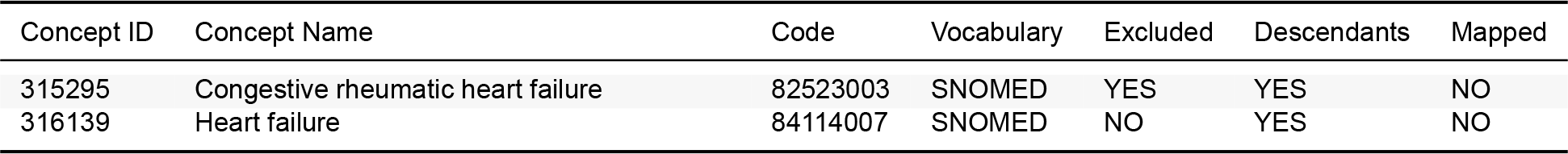

#### B.2 4-point MACE

##### B.2.1 Cohort Entry Events

People may enter the cohort when observing any of the following:

1. condition occurrences of ‘Acute myocardial Infarction’.
2. condition occurrences of ‘Sudden cardiac death’.
3. condition occurrences of ‘Ischemic stroke’.
4. condition occurrences of ‘Iintracranial bleed Hemorrhagic stroke’.
5. condition occurrences of ‘Heart Failure’.

Restrict entry events to having at least 1 visit occurrence of ‘Inpatient or ER visit’, starting anytime on or before cohort entry start date and ending between 0 days before and all days after cohort entry start date.

##### B.2.2 Cohort Exit

The cohort end date will be offset from index event’s start date plus 7 days.

##### B.2.3 Cohort Eras

Entry events will be combined into cohort eras if they are within 180 days of each other.

##### B.2.4 Concept: Inpatient or ER visit

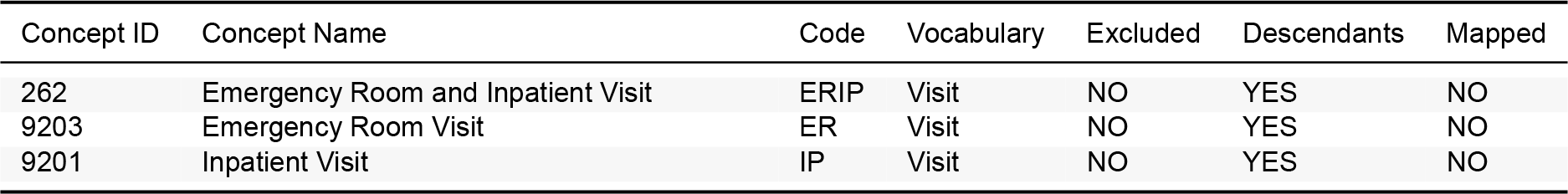

##### B.2.5 Concept: Acute myocardial Infarction

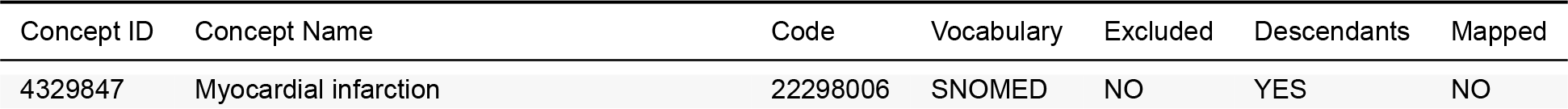
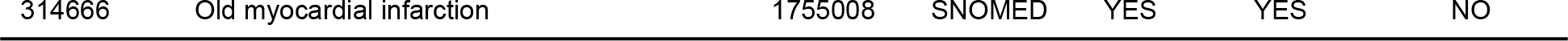

| Concept ID | Concept Name | Code | Vocabulary | Excluded | Descendants | Mapped |
| --- | --- | --- | --- | --- | --- | --- |
| 4329847 | Myocardial infarction | 22298006 | SNOMED | NO | YES | NO |
| 314666 | Old myocardial infarction | 1755008 | SNOMED | YES | YES | NO |

##### B.2.6 Concept: Sudden cardiac death

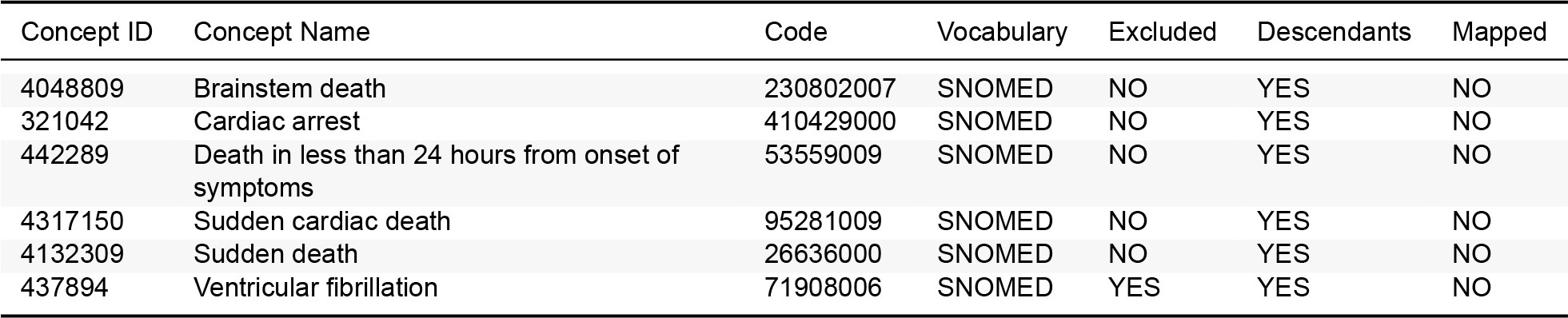

##### B.2.7 Concept: Ischemic stroke

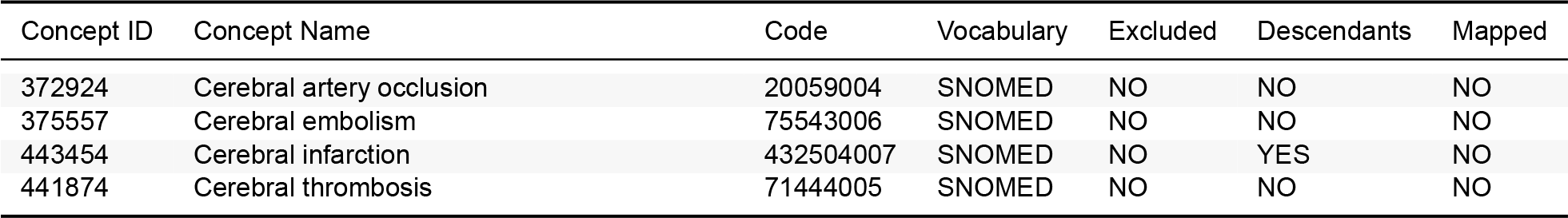

##### B.2.8 Concept: Iintracranial bleed Hemorrhagic stroke

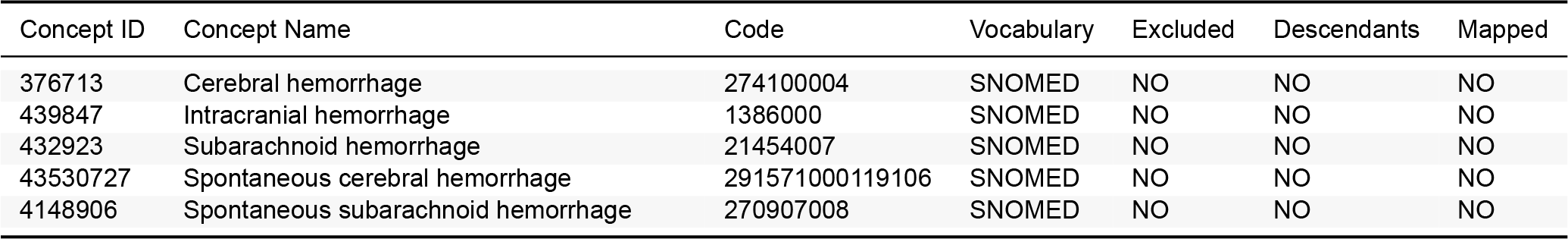

##### B.2.9 Concept: Heart Failure

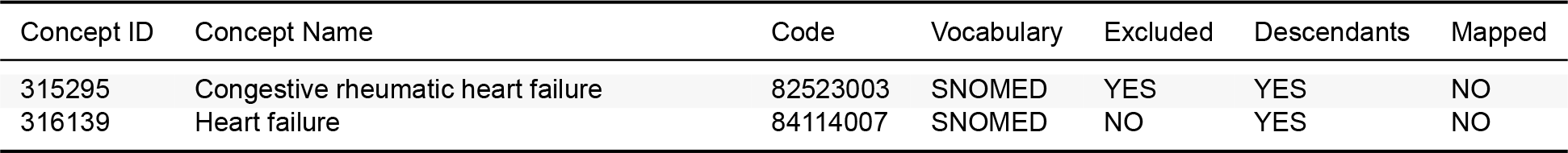

#### B.3 Acute myocardial infarction

##### B.3.1 Cohort Entry Events

People may enter the cohort when observing any of the following:

1. condition occurrences of ‘[LEGEND-T2DM] Acute myocardial Infarction’.

Restrict entry events to having at least 1 visit occurrence of ‘Inpatient or ER visit’, starting anytime on or before cohort entry start date and ending between 0 days before and all days after cohort entry start date.

##### B.3.2 Cohort Exit

The cohort end date will be offset from index event’s start date plus 7 days.

##### B.3.3 Cohort Eras

Entry events will be combined into cohort eras if they are within 180 days of each other.

##### B.3.4 Concept: Inpatient or ER visit

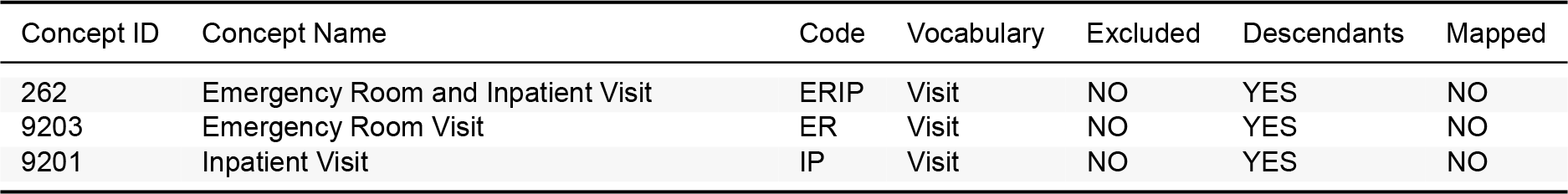

##### B.3.5 Concept: [LEGEND-T2DM] Acute myocardial Infarction

| Concept ID | Concept Name | Code | Vocabulary | Excluded | Descendants | Mapped |
| --- | --- | --- | --- | --- | --- | --- |
| 4329847 | Myocardial infarction | 22298006 | SNOMED | NO | YES | NO |
| 314666 | Old myocardial infarction | 1755008 | SNOMED | YES | YES | NO |

#### B.4 Acute renal failure

##### B.4.1 Cohort Entry Events

People may enter the cohort when observing any of the following:

1. condition occurrences of ‘Acute Renal Failure’.

Restrict entry events to having at least 1 visit occurrence of ‘Inpatient or ER visit’, starting anytime on or before cohort entry start date and ending between 0 days before and all days after cohort entry start date.

##### B.4.2 Cohort Exit

The cohort end date will be offset from index event’s start date plus 30 days.

##### B.4.3 Cohort Eras

Entry events will be combined into cohort eras if they are within 30 days of each other.

##### B.4.4 Concept: Inpatient or ER visit

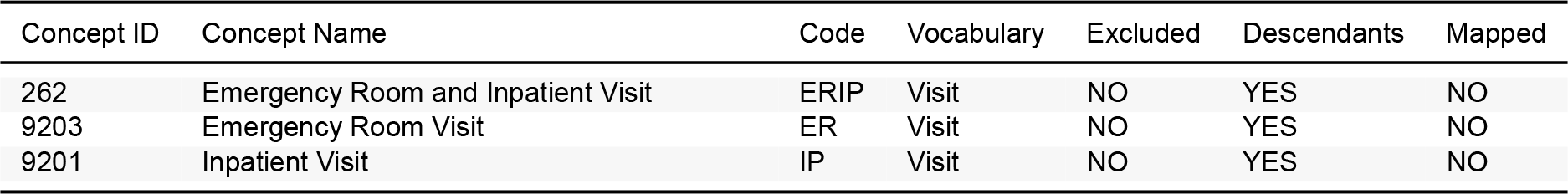

##### B.4.5 Concept: Acute Renal Failure

| Concept ID | Concept Name | Code | Vocabulary | Excluded | Descendants | Mapped |
| --- | --- | --- | --- | --- | --- | --- |
| 197320 | Acute renal failure syndrome | 14669001 | SNOMED | NO | YES | NO |
| 432961 | Acute renal papillary necrosis with renal failure | 298015003 | SNOMED | NO | YES | NO |
| 444044 | Acute tubular necrosis | 35455006 | SNOMED | NO | YES | NO |

#### B.5 Glycemic control

##### B.5.1 Cohort Entry Events

People enter the cohort when observing any of the following:

1. measurements of ‘HbA1c_v2’, numeric value <= 7; unit: “percent”.
2. measurements of ‘HbA1c_v2’, numeric value <= 53; unit: “millimole per mole”.

Limit qualifying entry events to the earliest event per person.

##### B.5.2 Cohort Exit

The cohort end date will be offset from index event’s start date plus 1 day.

##### B.5.3 Cohort Eras

Entry events will be combined into cohort eras if they are within 90 days of each other.

##### B.5.4 Concept: HbA1c_v2

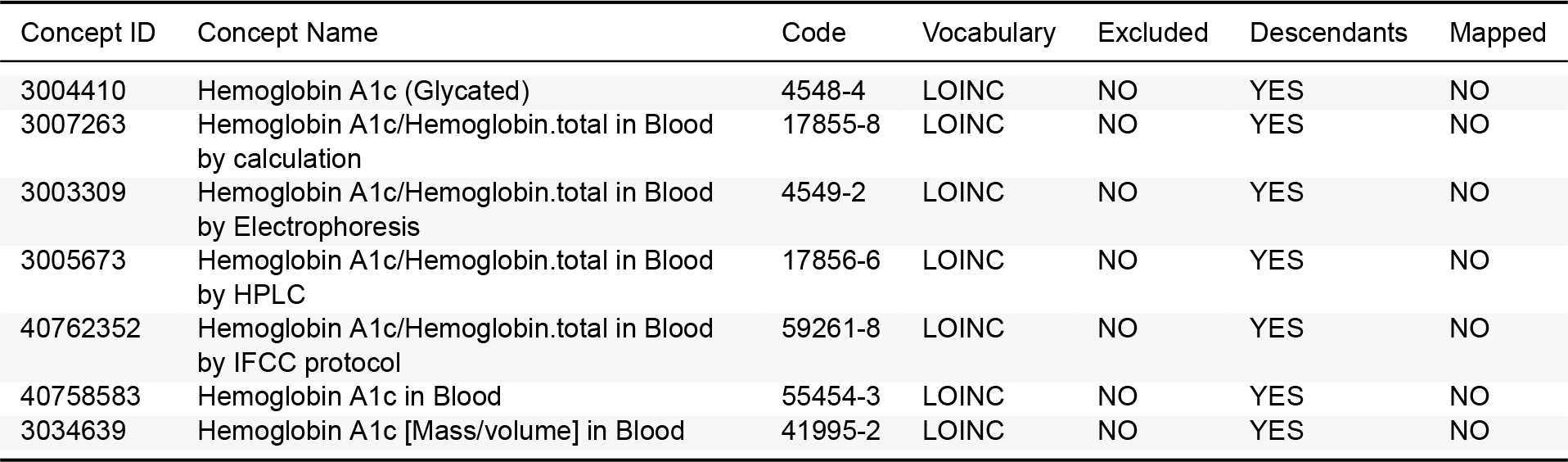

#### B.6 Hospitalization with heart failure

##### B.6.1 Cohort Entry Events

People enter the cohort when observing any of the following:

1. visit occurrences of ‘Inpatient or ER visit’; having at least 1 condition occurrence of ‘[LEGEND-T2DM] Heart Failure’, starting between 0 days before and all days after ‘Inpatient or ER visit’ start date and starting anytime on or before ‘Inpatient or ER visit’ end date.

##### B.6.2 Cohort Exit

The cohort end date will be offset from index event’s end date plus 0 days.

##### B.6.3 Cohort Eras

Entry events will be combined into cohort eras if they are within 7 days of each other.

##### B.6.4 Concept: Inpatient or ER visit

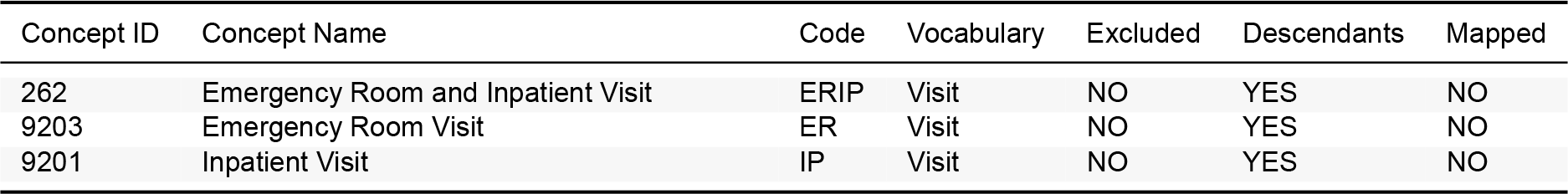

##### B.6.5 Concept: [LEGEND-T2DM] Heart Failure

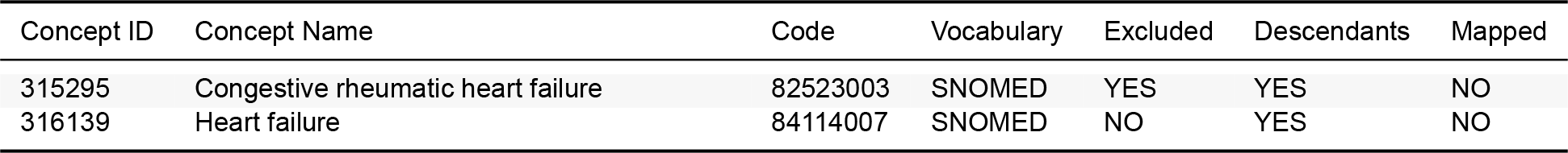

#### B.7 Measured renal dysfunction

##### B.7.1 Cohort Entry Events

People enter the cohort when observing any of the following:

1. measurements of ‘Creatinine measurement’, numeric value > 3; unit: “milligram per deciliter”.
2. measurements of ‘Creatinine measurement’, numeric value > 265; unit: “micromole/liter”.
3. measurements of ‘Creatinine measurement’, numeric value > 0.265; unit: “millimole per liter”.
4. measurements of ‘Creatinine measurement’, numeric value > 3; unit: “milligram”.

Limit cohort entry events to the earliest event per person.

##### B.7.2 Cohort Exit

The cohort end date will be offset from index event’s start date plus 1 day.

##### B.7.3 Cohort Eras

Entry events will be combined into cohort eras if they are within 0 days of each other.

##### B.7.4 Concept: Creatinine measurement

| Concept ID | Concept Name | Code | Vocabulary | Excluded | Descendants | Mapped |
| --- | --- | --- | --- | --- | --- | --- |
| 3016723 | Creatinine [Mass/volume] in Serum or Plasma | 2160-0 | LOINC | NO | YES | NO |
| 3022243 | Creatinine [Mass/volume] in Serum or Plasma –pre dialysis | 11042-9 | LOINC | NO | YES | NO |
| 3020564 | Creatinine [Moles/volume] in Serum or Plasma | 14682-9 | LOINC | NO | YES | NO |

#### B.8 Revascularization

##### B.8.1 Cohort Entry Events

People may enter the cohort when observing any of the following:

1. procedure occurrences of ‘PCI’.
2. procedure occurrences of ‘CABG’.

##### B.8.2 Additional Inclusion Criteria

- Hospitalization

Entry events having at least 1 visit occurrence of ‘Hospitalization’, starting between 0 days before and 0 days after cohort entry start date.

##### B.8.3 Cohort Exit

The cohort end date will be offset from index event’s start date plus 1 day.

##### B.8.4 Cohort Eras

Entry events will be combined into cohort eras if they are within 0 days of each other.

##### B.8.5 Concept: PCI

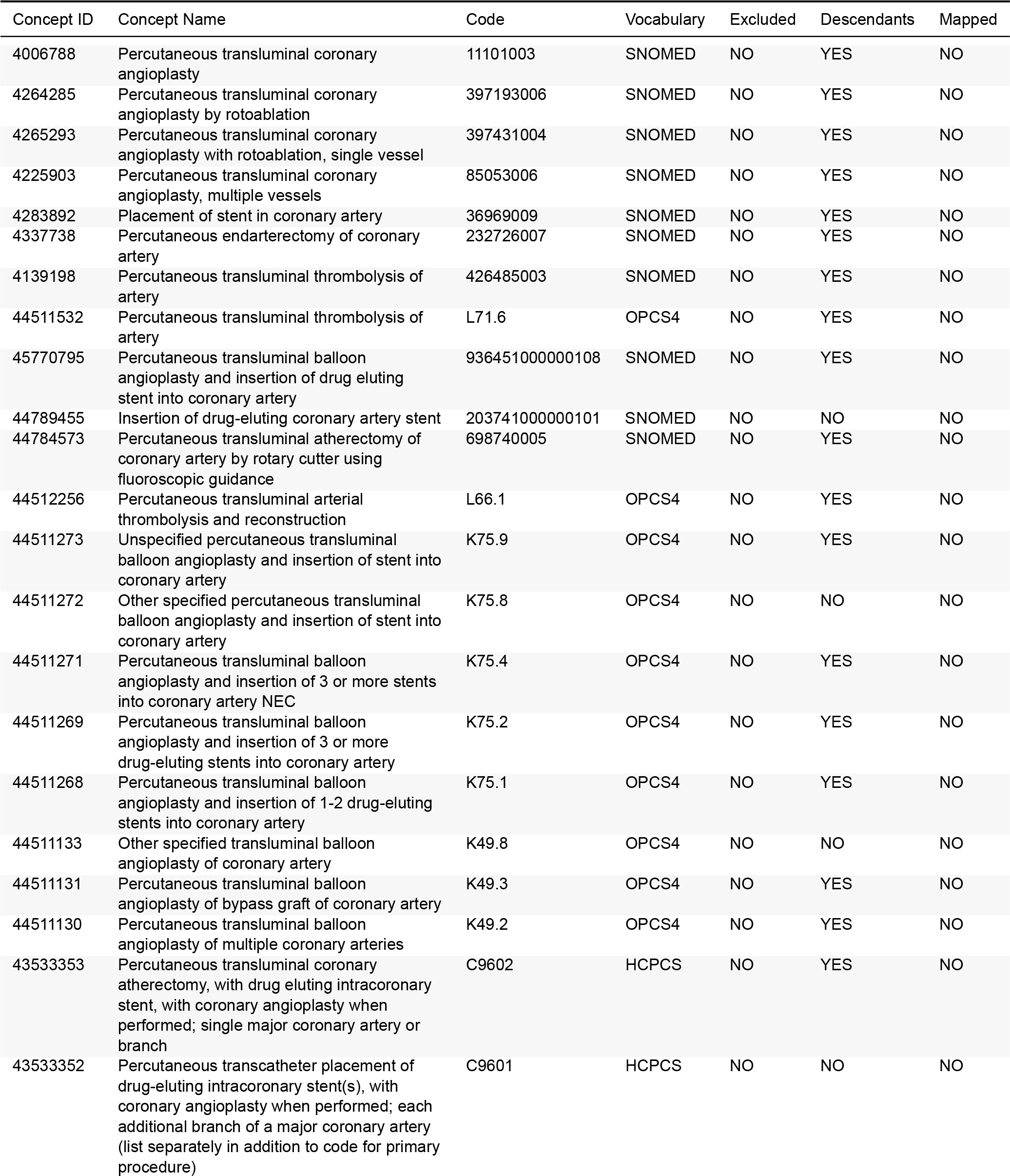
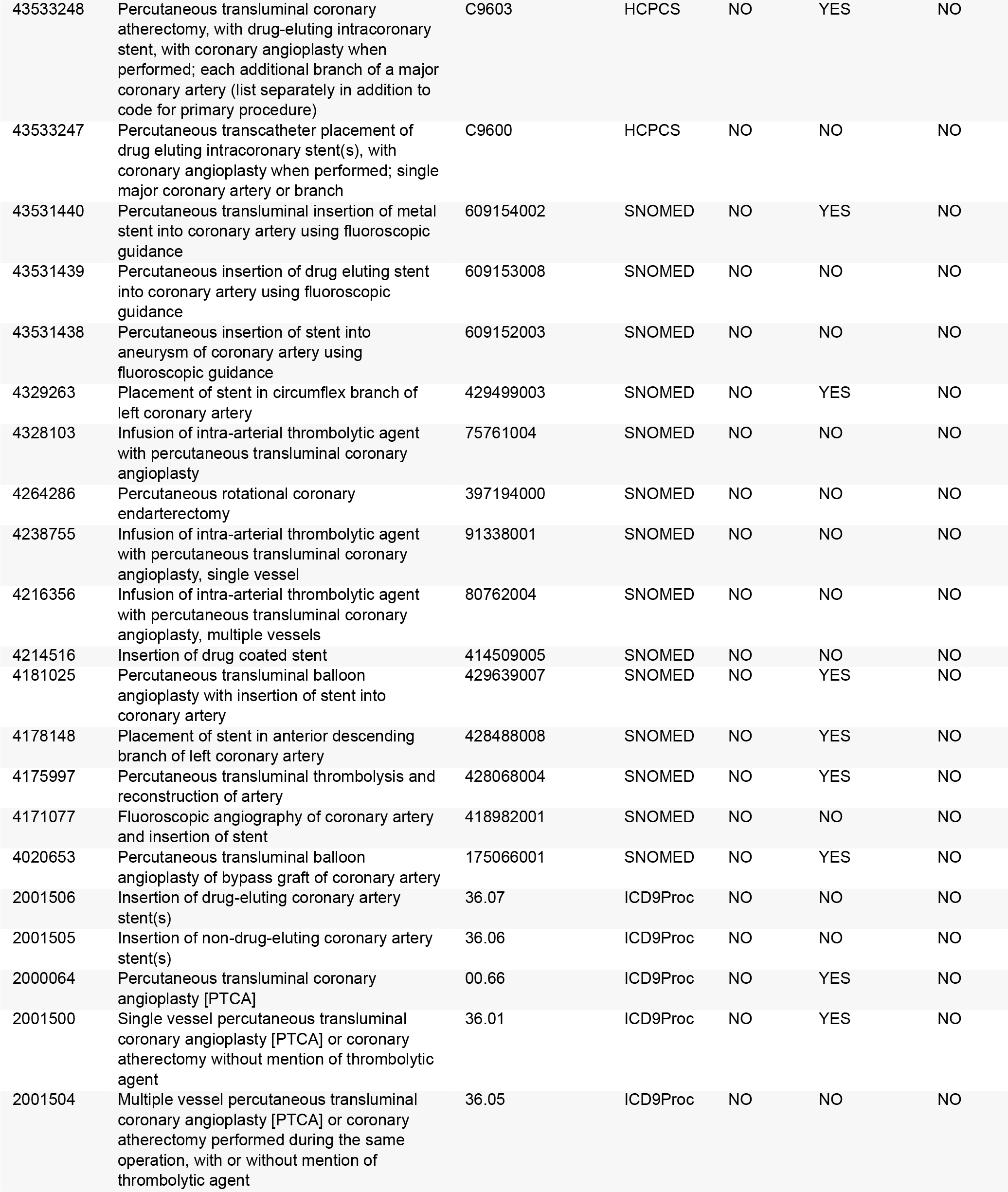
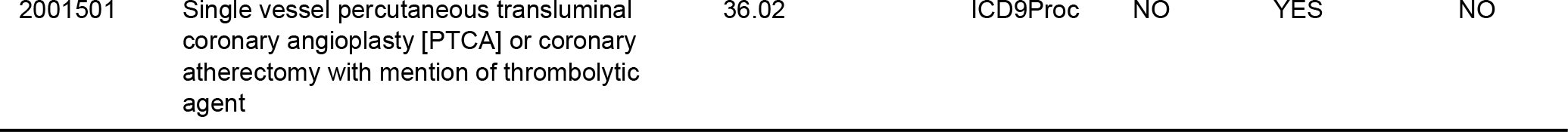

| Concept ID | Concept Name | Code | Vocabulary | Excluded | Descendants | Mapped |
| --- | --- | --- | --- | --- | --- | --- |
| 4006788 | Percutaneous transluminal coronary angioplasty | 11101003 | SNOMED | NO | YES | NO |
| 4264285 | Percutaneous transluminal coronary angioplasty by rotoablation | 397193006 | SNOMED | NO | YES | NO |
| 4265293 | Percutaneous transluminal coronary angioplasty with rotoablation, single vessel | 397431004 | SNOMED | NO | YES | NO |
| 4225903 | Percutaneous transluminal coronary angioplasty, multiple vessels | 85053006 | SNOMED | NO | YES | NO |
| 4283892 | Placement of stent in coronary artery | 36969009 | SNOMED | NO | YES | NO |
| 4337738 | Percutaneous endarterectomy of coronary artery | 232726007 | SNOMED | NO | YES | NO |
| 4139198 | Percutaneous transluminal thrombolysis of artery | 426485003 | SNOMED | NO | YES | NO |
| 44511532 | Percutaneous transluminal thrombolysis of artery | L71.6 | OPCS4 | NO | YES | NO |
| 45770795 | Percutaneous transluminal balloon angioplasty and insertion of drug eluting stent into coronary artery | 936451000000108 | SNOMED | NO | YES | NO |
| 44789455 | Insertion of drug-eluting coronary artery stent | 203741000000101 | SNOMED | NO | NO | NO |
| 44784573 | Percutaneous transluminal atherectomy of coronary artery by rotary cutter using fluoroscopic guidance | 698740005 | SNOMED | NO | YES | NO |
| 44512256 | Percutaneous transluminal arterial thrombolysis and reconstruction | L66.1 | OPCS4 | NO | YES | NO |
| 44511273 | Unspecified percutaneous transluminal balloon angioplasty and insertion of stent into coronary artery | K75.9 | OPCS4 | NO | YES | NO |
| 44511272 | Other specified percutaneous transluminal balloon angioplasty and insertion of stent into coronary artery | K75.8 | OPCS4 | NO | NO | NO |
| 44511271 | Percutaneous transluminal balloon angioplasty and insertion of 3 or more stents into coronary artery NEC | K75.4 | OPCS4 | NO | YES | NO |
| 44511269 | Percutaneous transluminal balloon angioplasty and insertion of 3 or more drug-eluting stents into coronary artery | K75.2 | OPCS4 | NO | YES | NO |
| 44511268 | Percutaneous transluminal balloon angioplasty and insertion of 1-2 drug-eluting stents into coronary artery | K75.1 | OPCS4 | NO | YES | NO |
| 44511133 | Other specified transluminal balloon angioplasty of coronary artery | K49.8 | OPCS4 | NO | NO | NO |
| 44511131 | Percutaneous transluminal balloon angioplasty of bypass graft of coronary artery | K49.3 | OPCS4 | NO | YES | NO |
| 44511130 | Percutaneous transluminal balloon angioplasty of multiple coronary arteries | K49.2 | OPCS4 | NO | YES | NO |
| 43533353 | Percutaneous transluminal coronary atherectomy, with drug eluting intracoronary stent, with coronary angioplasty when performed; single major coronary artery or branch | C9602 | HCPCS | NO | YES | NO |
| 43533352 | Percutaneous transcatheter placement of drug-eluting intracoronary stent(s), with coronary angioplasty when performed; each additional branch of a major coronary artery (list separately in addition to code for primary procedure) | C9601 | HCPCS | NO | NO | NO |
| 43533248 | Percutaneous transluminal coronary atherectomy, with drug-eluting intracoronary stent, with coronary angioplasty when performed; each additional branch of a major coronary artery (list separately in addition to code for primary procedure) | C9603 | HCPCS | NO | YES | NO |
| 43533247 | Percutaneous transcatheter placement of drug eluting intracoronary stent(s), with coronary angioplasty when performed; single major coronary artery or branch | C9600 | HCPCS | NO | NO | NO |
| 43531440 | Percutaneous transluminal insertion of metal stent into coronary artery using fluoroscopic guidance | 609154002 | SNOMED | NO | YES | NO |
| 43531439 | Percutaneous insertion of drug eluting stent into coronary artery using fluoroscopic guidance | 609153008 | SNOMED | NO | NO | NO |
| 43531438 | Percutaneous insertion of stent into aneurysm of coronary artery using fluoroscopic guidance | 609152003 | SNOMED | NO | NO | NO |
| 4329263 | Placement of stent in circumflex branch of left coronary artery | 429499003 | SNOMED | NO | YES | NO |
| 4328103 | Infusion of intra-arterial thrombolytic agent with percutaneous transluminal coronary angioplasty | 75761004 | SNOMED | NO | NO | NO |
| 4264286 | Percutaneous rotational coronary endarterectomy | 397194000 | SNOMED | NO | NO | NO |
| 4238755 | Infusion of intra-arterial thrombolytic agent with percutaneous transluminal coronary angioplasty, single vessel | 91338001 | SNOMED | NO | NO | NO |
| 4216356 | Infusion of intra-arterial thrombolytic agent with percutaneous transluminal coronary angioplasty, multiple vessels | 80762004 | SNOMED | NO | NO | NO |
| 4214516 | Insertion of drug coated stent | 414509005 | SNOMED | NO | NO | NO |
| 4181025 | Percutaneous transluminal balloon angioplasty with insertion of stent into coronary artery | 429639007 | SNOMED | NO | YES | NO |
| 4178148 | Placement of stent in anterior descending branch of left coronary artery | 428488008 | SNOMED | NO | YES | NO |
| 4175997 | Percutaneous transluminal thrombolysis and reconstruction of artery | 428068004 | SNOMED | NO | YES | NO |
| 4171077 | Fluoroscopic angiography of coronary artery and insertion of stent | 418982001 | SNOMED | NO | NO | NO |
| 4020653 | Percutaneous transluminal balloon angioplasty of bypass graft of coronary artery | 175066001 | SNOMED | NO | YES | NO |
| 2001506 | Insertion of drug-eluting coronary artery stent(s) | 36.07 | ICD9Proc | NO | NO | NO |
| 2001505 | Insertion of non-drug-eluting coronary artery stent(s) | 36.06 | ICD9Proc | NO | NO | NO |
| 2000064 | Percutaneous transluminal coronary angioplasty [PTCA] | 00.66 | ICD9Proc | NO | YES | NO |
| 2001500 | Single vessel percutaneous transluminal coronary angioplasty [PTCA] or coronary atherectomy without mention of thrombolytic agent | 36.01 | ICD9Proc | NO | YES | NO |
| 2001504 | Multiple vessel percutaneous transluminal coronary angioplasty [PTCA] or coronary atherectomy performed during the same operation, with or without mention of thrombolytic agent | 36.05 | ICD9Proc | NO | NO | NO |
| 2001501 | Single vessel percutaneous transluminal coronary angioplasty [PTCA] or coronary atherectomy with mention of thrombolytic agent | 36.02 | ICD9Proc | NO | YES | NO |

##### B.8.6 Concept: Hospitalization

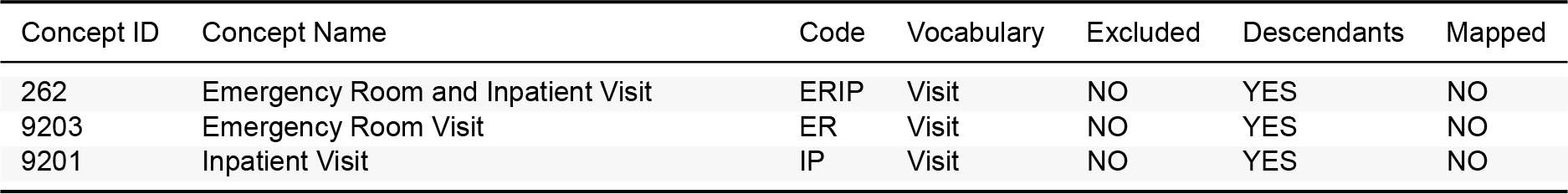

##### B.8.7 Concept: CABG

| Concept ID | Concept Name | Code | Vocabulary | Excluded | Descendants | Mapped |
| --- | --- | --- | --- | --- | --- | --- |
| 2001516 | Abdominal-coronary artery bypass | 36.17 | ICD9Proc | NO | YES | NO |
| 4284104 | Aortocoronary artery bypass graft | 67166004.00 | SNOMED | NO | YES | NO |
| 4229433 | Aortocoronary artery bypass graft with prosthesis | 8876004.00 | SNOMED | NO | YES | NO |
| 4146972 | Aortocoronary artery bypass graft with saphenous vein graft | 3546002.00 | SNOMED | NO | YES | NO |
| 4228305 | Aortocoronary artery bypass graft with three vein grafts | 405599002.00 | SNOMED | NO | YES | NO |
| 4228304 | Aortocoronary artery bypass graft with two vein grafts | 405598005.00 | SNOMED | NO | YES | NO |
| 4063237 | Aortocoronary artery bypass graft with vein graft | 17073005.00 | SNOMED | NO | YES | NO |
| 4148030 | Aortocoronary bypass grafting | 309814006.00 | SNOMED | NO | YES | NO |
| 4008625 | Aortocoronary bypass of four or more coronary arteries | 10190003.00 | SNOMED | NO | YES | NO |
| 4106548 | Aortocoronary bypass of one coronary artery | 29819009.00 | SNOMED | NO | YES | NO |
| 4031996 | Aortocoronary bypass of three coronary arteries | 14323007.00 | SNOMED | NO | YES | NO |
| 4234990 | Aortocoronary bypass of two coronary arteries | 90487008.00 | SNOMED | NO | YES | NO |
| 45889469 | Arterial Grafting for Coronary Artery Bypass | 1006216.00 | CPT4 | NO | YES | NO |
| 4240486 | Carotid-subclavian artery bypass graft with vein | 59012002.00 | SNOMED | NO | YES | NO |
| 4336464 | Coronary artery bypass graft | 232717009.00 | SNOMED | NO | YES | NO |
| 4337056 | Coronary artery bypass graft x 1 | 232719007.00 | SNOMED | NO | YES | NO |
| 4000733 | Coronary artery bypass graft, anastomosis of artery of thorax to coronary artery | 119565001.00 | SNOMED | NO | YES | NO |
| 4336467 | Coronary artery bypass grafts greater than 5 | 232724005.00 | SNOMED | NO | YES | NO |
| 4336465 | Coronary artery bypass grafts x 2 | 232720001.00 | SNOMED | NO | YES | NO |
| 4339629 | Coronary artery bypass grafts x 3 | 232721002.00 | SNOMED | NO | YES | NO |
| 4337737 | Coronary artery bypass grafts x 4 | 232722009.00 | SNOMED | NO | YES | NO |
| 4336466 | Coronary artery bypass grafts x 5 | 232723004.00 | SNOMED | NO | YES | NO |
| 4233421 | Coronary artery bypass with autogenous graft of internal mammary artery, single graft | 359601003.00 | SNOMED | NO | YES | NO |
| 4305509 | Coronary artery bypass with autogenous graft, five grafts | 82247006.00 | SNOMED | NO | YES | NO |
| 4309432 | Coronary artery bypass with autogenous graft, four grafts | 39202005.00 | SNOMED | NO | YES | NO |
| 4011931 | Coronary artery bypass with autogenous graft, three grafts | 10326007.00 | SNOMED | NO | YES | NO |
| 4253805 | Coronary artery bypass with autogenous graft, two grafts | 74371005.00 | SNOMED | NO | YES | NO |
| 45887879 | Coronary artery bypass, using arterial graft(s) | 1006217.00 | CPT4 | NO | YES | NO |
| 2107242 | Coronary artery bypass, using arterial graft(s); 2 coronary arterial grafts | 33534.00 | CPT4 | NO | YES | NO |
| 2107243 | Coronary artery bypass, using arterial graft(s); 3 coronary arterial grafts | 33535.00 | CPT4 | NO | YES | NO |
| 2107244 | Coronary artery bypass, using arterial graft(s); 4 or more coronary arterial grafts | 33536.00 | CPT4 | NO | YES | NO |
| 2107231 | Coronary artery bypass, using arterial graft(s); single arterial graft | 33533.00 | CPT4 | NO | YES | NO |
| 45889898 | Coronary artery bypass, using venous graft(s) and arterial graft(s) | 1006208.00 | CPT4 | NO | YES | NO |
| 2107223 | Coronary artery bypass, using venous graft(s) and arterial graft(s); 2 venous grafts (List separately in addition to code for primary procedure) | 33518.00 | CPT4 | NO | YES | NO |
| 2107224 | Coronary artery bypass, using venous graft(s) and arterial graft(s); 3 venous grafts (List separately in addition to code for primary procedure) | 33519.00 | CPT4 | NO | YES | NO |
| 2107226 | Coronary artery bypass, using venous graft(s) and arterial graft(s); 4 venous grafts (List separately in addition to code for primary procedure) | 33521.00 | CPT4 | NO | YES | NO |
| 2107227 | Coronary artery bypass, using venous graft(s) and arterial graft(s); 5 venous grafts (List separately in addition to code for primary procedure) | 33522.00 | CPT4 | NO | YES | NO |
| 2107228 | Coronary artery bypass, using venous graft(s) and arterial graft(s); 6 or more venous grafts (List separately in addition to code for primary procedure) | 33523.00 | CPT4 | NO | YES | NO |
| 2107222 | Coronary artery bypass, using venous graft(s) and arterial graft(s); single vein graft (List separately in addition to code for primary procedure) | 33517.00 | CPT4 | NO | YES | NO |
| 45887862 | Coronary artery bypass, vein only | 1006200.00 | CPT4 | NO | YES | NO |
| 2107217 | Coronary artery bypass, vein only; 2 coronary venous grafts | 33511.00 | CPT4 | NO | YES | NO |
| 2107218 | Coronary artery bypass, vein only; 3 coronary venous grafts | 33512.00 | CPT4 | NO | YES | NO |
| 2107219 | Coronary artery bypass, vein only; 4 coronary venous grafts | 33513.00 | CPT4 | NO | YES | NO |
| 2107220 | Coronary artery bypass, vein only; 5 coronary venous grafts | 33514.00 | CPT4 | NO | YES | NO |
| 2107221 | Coronary artery bypass, vein only; 6 or more coronary venous grafts | 33516.00 | CPT4 | NO | YES | NO |
| 2107216 | Coronary artery bypass, vein only; single coronary venous graft | 33510.00 | CPT4 | NO | YES | NO |
| 2001515 | Double internal mammary-coronary artery bypass | 36.16 | ICD9Proc | NO | YES | NO |
| 4000732 | Internal mammary-coronary artery bypass graft | 119564002.00 | SNOMED | NO | YES | NO |
| 2001514 | Single internal mammary-coronary artery bypass | 36.15 | ICD9Proc | NO | YES | NO |
| 4233420 | Single internal mammary-coronary artery bypass | 359597003.00 | SNOMED | NO | YES | NO |
| 45889467 | Venous Grafting Only for Coronary Artery Bypass | 1006199.00 | CPT4 | NO | YES | NO |
| 4020216 | Revision of bypass for coronary artery | 175036008.00 | SNOMED | NO | NO | NO |
| 4305852 | Off-pump coronary artery bypass | 418824004.00 | SNOMED | NO | NO | NO |

#### B.9 Stroke

##### B.9.1 Cohort Entry Events

People may enter the cohort when observing any of the following:

1. condition occurrences of ‘[LEGEND-T2DM] Stroke (ischemic or hemorrhagic)’.

Restrict entry events to having at least 1 visit occurrence of ‘Inpatient or ER visit’, starting between all days before and 1 days after cohort entry start date and ending between 0 days before and all days after cohort entry start date.

##### B.9.2 Cohort Exit

The cohort end date will be offset from index event’s start date plus 7 days.

##### B.9.3 Cohort Eras

Entry events will be combined into cohort eras if they are within 180 days of each other.

##### B.9.4 Concept: Inpatient or ER visit

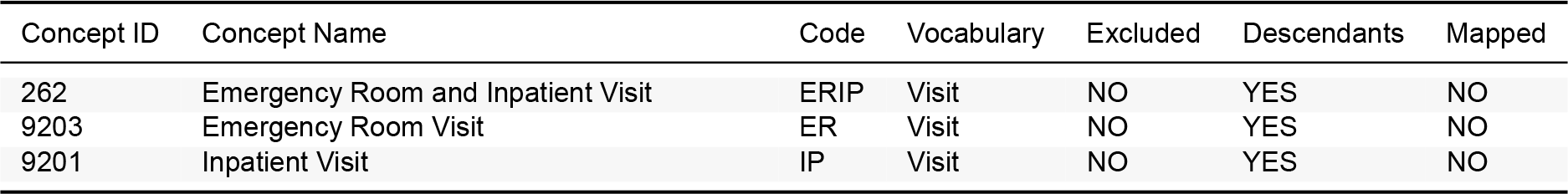

##### B.9.5 Concept: [LEGEND-T2DM] Stroke (ischemic or hemorrhagic)

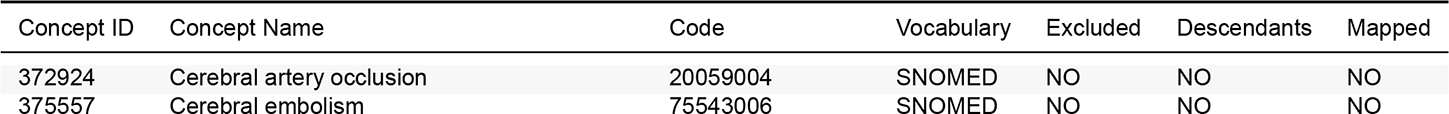
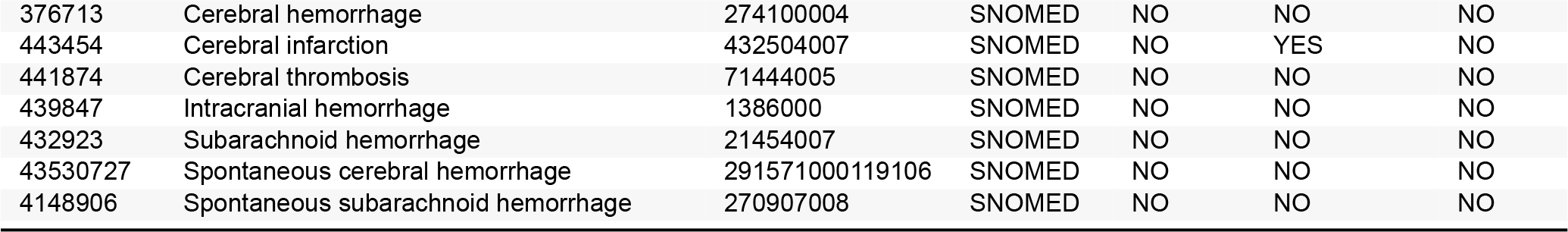

| Concept ID | Concept Name | Code | Vocabulary | Excluded | Descendants | Mapped |
| --- | --- | --- | --- | --- | --- | --- |
| 372924 | Cerebral artery occlusion | 20059004 | SNOMED | NO | NO | NO |
| 375557 | Cerebral embolism | 75543006 | SNOMED | NO | NO | NO |
| 376713 | Cerebral hemorrhage | 274100004 | SNOMED | NO | NO | NO |
| 443454 | Cerebral infarction | 432504007 | SNOMED | NO | YES | NO |
| 441874 | Cerebral thrombosis | 71444005 | SNOMED | NO | NO | NO |
| 439847 | Intracranial hemorrhage | 1386000 | SNOMED | NO | NO | NO |
| 432923 | Subarachnoid hemorrhage | 21454007 | SNOMED | NO | NO | NO |
| 43530727 | Spontaneous cerebral hemorrhage | 291571000119106 | SNOMED | NO | NO | NO |
| 4148906 | Spontaneous subarachnoid hemorrhage | 270907008 | SNOMED | NO | NO | NO |

#### B.10 Sudden cardiac death

##### B.10.1 Cohort Entry Events

People may enter the cohort when observing any of the following:

1. condition occurrences of ‘[LEGEND HTN] Sudden cardiac death’.

Restrict entry events to having at least 1 visit occurrence of ‘Inpatient or ER visit’, starting anytime on or before cohort entry start date and ending between 0 days before and all days after cohort entry start date.

##### B.10.2 Cohort Exit

The cohort end date will be offset from index event’s start date plus 7 days.

##### B.10.3 Cohort Eras

Entry events will be combined into cohort eras if they are within 180 days of each other.

##### B.10.4 Concept: Inpatient or ER visit

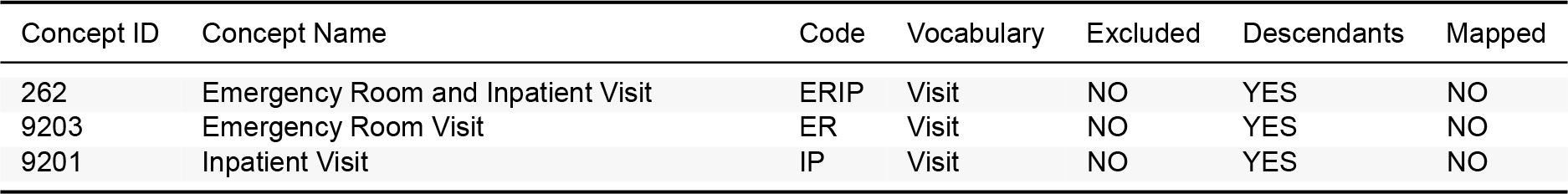

##### B.10.5 Concept: [LEGEND HTN] Sudden cardiac death

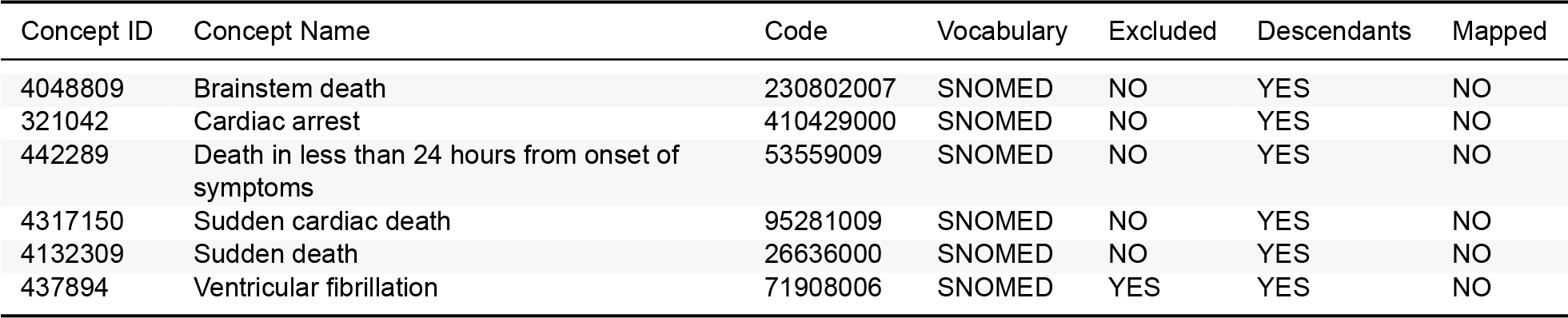

#### B.11 Abnormal weight gain

##### B.11.1 Cohort Entry Events

People enter the cohort when observing any of the following:

1. observations of ‘[LEGEND HTN] Abnormal weight gain’.

##### B.11.2 Cohort Exit

The cohort end date will be offset from index event’s start date plus 1 day.

##### B.11.3 Cohort Eras

Entry events will be combined into cohort eras if they are within 90 days of each other.

##### B.11.4 Concept: [LEGEND HTN] Abnormal weight gain

| Concept ID | Concept Name | Code | Vocabulary | Excluded | Descendants | Mapped |
| --- | --- | --- | --- | --- | --- | --- |
| 439141 | Abnormal weight gain | 161833006 | SNOMED | NO | YES | NO |

#### B.12 Abnormal weight loss

##### B.12.1 Cohort Entry Events

People enter the cohort when observing any of the following:

1. observations of ‘[LEGEND HTN] Abnormal weight loss’.

##### B.12.2 Cohort Exit

The cohort end date will be offset from index event’s start date plus 1 day.

##### B.12.3 Cohort Eras

Entry events will be combined into cohort eras if they are within 90 days of each other.

##### B.12.4 Concept: [LEGEND HTN] Abnormal weight loss

| Concept ID | Concept Name | Code | Vocabulary | Excluded | Descendants | Mapped |
| --- | --- | --- | --- | --- | --- | --- |
| 435928 | Abnormal weight loss | 267024001 | SNOMED | NO | YES | NO |
| 40303297 | Weight loss (& abnormal) | 139091004 | SNOMED | NO | NO | NO |

#### B.13 Acute pancreatitis

##### B.13.1 Cohort Entry Events

People may enter the cohort when observing any of the following:

1. condition occurrences of ‘[LEGEND HTN] Acute pancreatitis’.

Restrict entry events to having at least 1 visit occurrence of ‘Inpatient or ER visit’, starting anytime on or before cohort entry start date and ending between 0 days before and all days after cohort entry start date.

##### B.13.2 Cohort Exit

The cohort end date will be offset from index event’s start date plus 7 days.

##### B.13.3 Cohort Eras

Entry events will be combined into cohort eras if they are within 30 days of each other.

##### B.13.4 Concept: Inpatient or ER visit

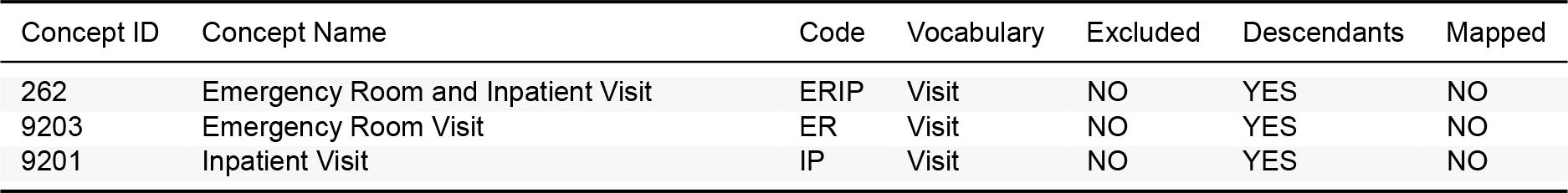

##### B.13.5 Concept: [LEGEND HTN] Acute pancreatitis

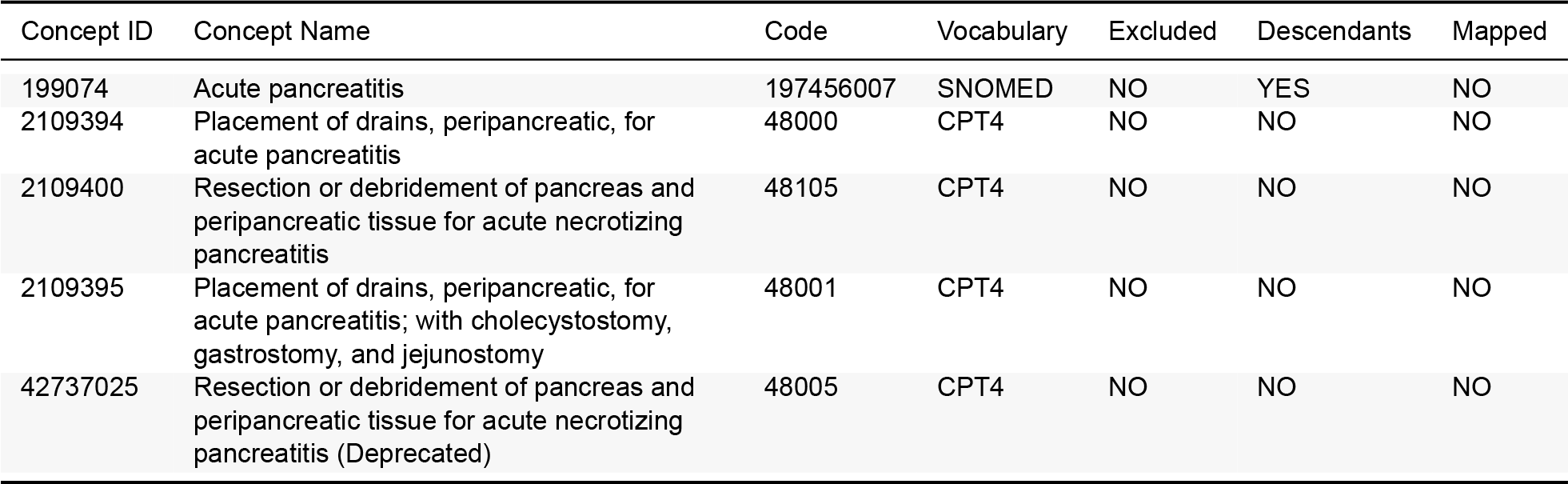

| Concept ID | Concept Name | Code | Vocabulary | Excluded | Descendants | Mapped |
| --- | --- | --- | --- | --- | --- | --- |
| 199074 | Acute pancreatitis | 197456007 | SNOMED | NO | YES | NO |
| 2109394 | Placement of drains, peripancreatic, for acute pancreatitis | 48000 | CPT4 | NO | NO | NO |
| 2109400 | Resection or debridement of pancreas and peripancreatic tissue for acute necrotizing pancreatitis | 48105 | CPT4 | NO | NO | NO |
| 2109395 | Placement of drains, peripancreatic, for acute pancreatitis; with cholecystostomy, gastrostomy, and jejunostomy | 48001 | CPT4 | NO | NO | NO |
| 42737025 | Resection or debridement of pancreas and peripancreatic tissue for acute necrotizing pancreatitis (Deprecated) | 48005 | CPT4 | NO | NO | NO |

#### B.14 All-cause mortality

##### B.14.1 Cohort Entry Events

People enter the cohort when observing any of the following:

1. death of any form.

Limit cohort entry events to the earliest event per person.

The person also exists the cohort at the end of continuous observation.

##### B.14.3 Cohort Eras

Entry events will be combined into cohort eras if they are within 0 days of each other.

#### B.15 Bladder cancer

##### B.15.1 Cohort Entry Events

People with continuous observation of 365 days before event enter the cohort when observing any of the following:

1. condition occurrence of ‘Bladder cancer’ for the first time in the person’s history.

Limit cohort entry events to the earliest event per person.

##### B.15.2 Cohort Exit

The person also exists the cohort at the end of continuous observation.

##### B.15.3 Cohort Eras

Entry events will be combined into cohort eras if they are within 0 days of each other.

##### B.15.4 Concept: Bladder cancer

| Concept ID | Concept Name | Code | Vocabulary | Excluded | Descendants | Mapped |
| --- | --- | --- | --- | --- | --- | --- |
| 197508 | Malignant tumor of urinary bladder | 399326009 | SNOMED | NO | YES | NO |

#### B.16 Bone fracture

##### B.16.1 Cohort Entry Events

People enter the cohort when observing any of the following:

1. condition occurrences of ‘Bone fracture’.

##### B.16.2 Cohort Exit

The cohort end date will be offset from index event’s start date plus 1 day.

##### B.16.3 Cohort Eras

Entry events will be combined into cohort eras if they are within 90 days of each other.

##### B.16.4 Concept: Bone fracture

| Concept ID | Concept Name | Code | Vocabulary | Excluded | Descendants | Mapped |
| --- | --- | --- | --- | --- | --- | --- |
| 75053 | Fracture of bone | 125605004 | SNOMED | NO | YES | NO |
| 4071354 | Open reduction of fracture with internal fixation | 20701002 | SNOMED | NO | YES | NO |

#### B.17 Breast cancer

##### B.17.1 Cohort Entry Events

People with continuous observation of 365 days before event enter the cohort when observing any of the following:

1. condition occurrence of ‘Malignant tumor of breast’ for the first time in the person’s history.

Limit cohort entry events to the earliest event per person.

##### B.17.2 Cohort Exit

The person also exists the cohort at the end of continuous observation.

##### B.17.3 Cohort Eras

Entry events will be combined into cohort eras if they are within 0 days of each other.

##### B.17.4 Concept: Malignant tumor of breast

| Concept ID | Concept Name | Code | Vocabulary | Excluded | Descendants | Mapped |
| --- | --- | --- | --- | --- | --- | --- |
| 4112853 | Malignant tumor of breast | 254837009 | SNOMED | NO | YES | NO |

##### B.18 Diabetic ketoacidosis

###### B.18.1 Cohort Entry Events

People may enter the cohort when observing any of the following:

1. condition occurrences of ‘Diabetic ketoacidosis’.

Restrict entry events to having at least 1 visit occurrence of ‘Inpatient or ER visit’, starting between all days before and 1 days after cohort entry start date and ending between 0 days before and all days after cohort entry start date.

###### B.18.2 Cohort Exit

The cohort end date will be offset from index event’s start date plus 7 days.

###### B.18.3 Cohort Eras

Entry events will be combined into cohort eras if they are within 180 days of each other.

###### B.18.4 Concept: Inpatient or ER visit

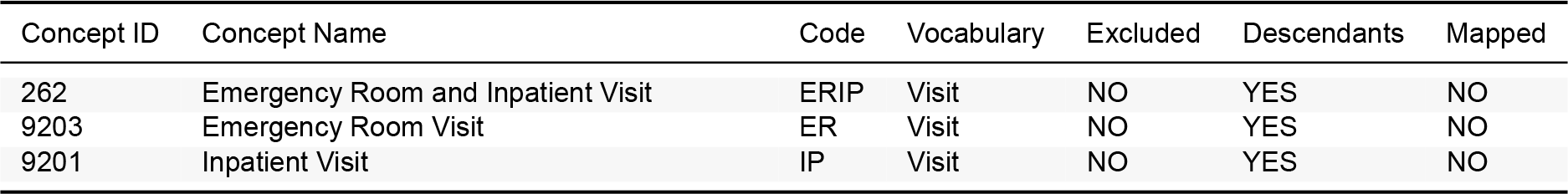

###### B.18.5 Concept: Diabetic ketoacidosis

| Concept ID | Concept Name | Code | Vocabulary | Excluded | Descendants | Mapped |
| --- | --- | --- | --- | --- | --- | --- |
| 443727 | Diabetic ketoacidosis | 420422005 | SNOMED | NO | YES | NO |

##### B.19 Diarrhea

###### B.19.1 Cohort Entry Events

People enter the cohort when observing any of the following:

1. condition occurrences of ‘[LEGEND HTN} Diarrhea’.

###### B.19.2 Cohort Exit

The cohort end date will be offset from index event’s start date plus 1 day.

###### B.19.3 Cohort Eras

Entry events will be combined into cohort eras if they are within 30 days of each other.

###### B.19.4 Concept: [LEGEND HTN} Diarrhea

| Concept ID | Concept Name | Code | Vocabulary | Excluded | Descendants | Mapped |
| --- | --- | --- | --- | --- | --- | --- |
| 196523 | Diarrhea | 62315008 | SNOMED | NO | YES | NO |
| 4134607 | Diarrheal disorder | 128333008 | SNOMED | NO | YES | NO |
| 201773 | Enteritis of small intestine | 64613007 | SNOMED | NO | NO | NO |
| 80141 | Functional diarrhea | 47812002 | SNOMED | NO | YES | NO |
| 4207688 | Infectious enteritis | 55184003 | SNOMED | NO | NO | NO |
| 4324838 | Noninfectious enteritis | 71207007 | SNOMED | NO | NO | NO |
| 197596 | Toxic gastroenteritis | 71583005 | SNOMED | NO | YES | NO |
| 196620 | Viral enteritis | 78420004 | SNOMED | NO | YES | NO |

##### B.20 Genitourinary infection

###### B.20.1 Cohort Entry Events

People enter the cohort when observing any of the following:

1. condition occurrences of ‘UTI’.

Limit qualifying entry events to the earliest event per person.

###### B.20.2 Cohort Exit

The cohort end date will be offset from index event’s start date plus 1 day.

###### B.20.3 Cohort Eras

Entry events will be combined into cohort eras if they are within 30 days of each other.

###### B.20.4 Concept: UTI

| Concept ID | Concept Name | Code | Vocabulary | Excluded | Descendants | Mapped |
| --- | --- | --- | --- | --- | --- | --- |
| 81902 | Urinary tract infectious disease | 6.85660050e+07 | SNOMED | NO | YES | NO |
| 4167328 | Pyuria | 4.80000100e+06 | SNOMED | NO | YES | NO |
| 77340 | Genitourinary tract infection in pregnancy | 2.67204006e+08 | SNOMED | NO | YES | NO |
| 4265485 | Bacteriuria | 6.13730060e+07 | SNOMED | NO | YES | NO |
| 4126297 | Chronic obstructive pyelonephritis | 2.36379002e+08 | SNOMED | NO | YES | NO |
| 195588 | Cystitis | 3.88220070e+07 | SNOMED | NO | YES | NO |
| 198806 | Abscess of prostate | 8.72500500e+06 | SNOMED | YES | YES | NO |
| 4126267 | Chronic radiation cystitis | 2.36629009e+08 | SNOMED | YES | YES | NO |
| 194997 | Prostatitis | 9.71300200e+06 | SNOMED | YES | NO | NO |
| 4077499 | Sterile pyuria | 2.75742001e+08 | SNOMED | YES | YES | NO |
| 442345 | Syphilis of kidney | 5.95300010e+07 | SNOMED | YES | YES | NO |
| 4062493 | Mumps nephritis | 1.71210060e+07 | SNOMED | YES | YES | NO |
| 45757237 | Diphtheria tubulointerstitial nephropathy | 1.08607100e+15 | SNOMED | YES | YES | NO |
| 36714969 | Asymptomatic bacteriuria | 7.20406004e+08 | SNOMED | YES | YES | NO |
| 195743 | Diphtheritic cystitis | 4.82780010e+07 | SNOMED | YES | YES | NO |
| 201353 | Irradiation cystitis | 1.12510000e+07 | SNOMED | YES | YES | NO |
| 4047937 | Neonatal urinary tract infection | 1.23010090e+07 | SNOMED | YES | YES | NO |
| 201792 | Nongonococcal urethritis | 8.46190010e+07 | SNOMED | YES | YES | NO |
| 4128384 | Non-infective cystitis | 2.36623005e+08 | SNOMED | YES | NO | NO |
| 78357 | Reactive arthritis triad | 6.72240070e+07 | SNOMED | YES | YES | NO |
| 195313 | Urethral abscess | 6.72770020e+07 | SNOMED | YES | YES | NO |
| 197919 | Urethral stricture due to infection | 8.03750020e+07 | SNOMED | YES | YES | NO |
| 439349 | Cystitis associated with another disorder | 1.97845000e+08 | SNOMED | YES | NO | NO |
| 4227291 | Hemorrhagic cystitis | 8.76960040e+07 | SNOMED | YES | NO | NO |
| 4060312 | Infections of urethra in pregnancy | 1.99206009e+08 | SNOMED | YES | NO | NO |
| 4127564 | Acute cystitis - culture-negative | 2.36624004e+08 | SNOMED | YES | YES | NO |
| 4126141 | Chronic cystitis - culture negative | 2.36626002e+08 | SNOMED | YES | NO | NO |
| 4127565 | Recurrent cystitis - culture-negative | 2.36625003e+08 | SNOMED | YES | YES | NO |
| 4207186 | Viral infection by site | 3.12130009e+08 | SNOMED | YES | YES | NO |
| 4207190 | Fungal infection by site | 3.12146001e+08 | SNOMED | YES | YES | NO |
| 434557 | Tuberculosis | 5.67170010e+07 | SNOMED | YES | YES | NO |
| 432251 | Disease caused by parasite | 1.73220070e+07 | SNOMED | YES | YES | NO |
| 36102152 | Protozoal infectious disorders | 1.00370720e+07 | MedDRA | YES | YES | NO |
| 433417 | Gonorrhea | 1.56280030e+07 | SNOMED | YES | YES | NO |
| 36102938 | Chlamydial infections | 1.00085610e+07 | MedDRA | YES | YES | NO |

##### B.21 Hyperkalemia

###### B.21.1 Cohort Entry Events

People enter the cohort when observing any of the following:

1. condition occurrences of ‘[LEGEND HTN] Hyperkalemia’.
2. measurements of ‘[LEGEND HTN] Potassium measurement’, numeric value > 5.6; unit: “millimole per liter”.

###### B.21.2 Cohort Exit

The cohort end date will be offset from index event’s start date plus 1 day.

###### B.21.3 Cohort Eras

Entry events will be combined into cohort eras if they are within 90 days of each other.

###### B.21.4 Concept: [LEGEND HTN] Hyperkalemia

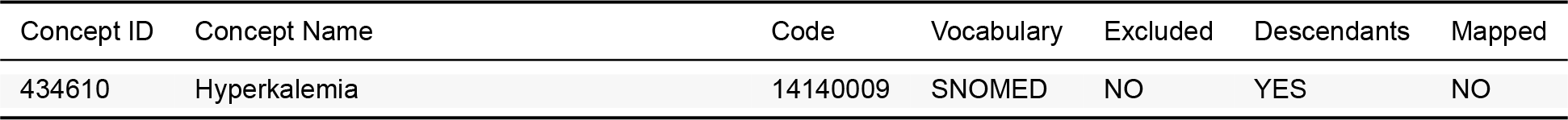

| Concept ID | Concept Name | Code | Vocabulary | Excluded | Descendants | Mapped |
| --- | --- | --- | --- | --- | --- | --- |
| 434610 | Hyperkalemia | 14140009 | SNOMED | NO | YES | NO |

###### B.21.5 Concept: [LEGEND HTN] Potassium measurement

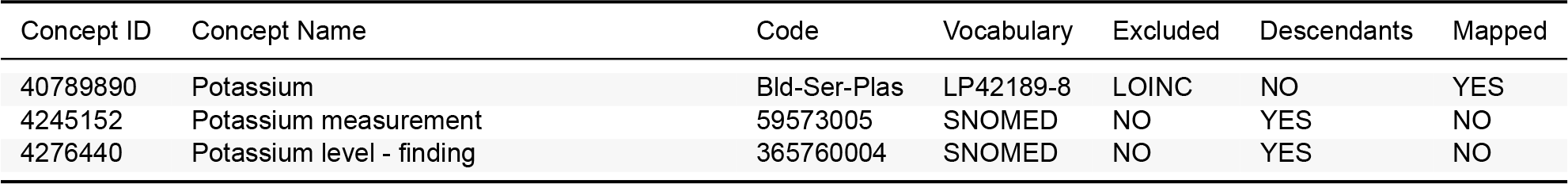

#### B.22 Hypoglycemia

##### B.22.1 Cohort Entry Events

People enter the cohort when observing any of the following:

1. condition occurrences of ‘Hypoglycemia’.

##### B.22.2 Cohort Exit

The cohort end date will be offset from index event’s start date plus 1 day.

##### B.22.3 Cohort Eras

Entry events will be combined into cohort eras if they are within 30 days of each other.

##### B.22.4 Concept: Hypoglycemia

| Concept ID | Concept Name | Code | Vocabulary | Excluded | Descendants | Mapped |
| --- | --- | --- | --- | --- | --- | --- |
| 380688 | Hypoglycemic coma | 267384006 | SNOMED | NO | YES | NO |
| 4048805 | Non-diabetic hypoglycemic coma | 230796005 | SNOMED | YES | YES | NO |
| 4226798 | Hypoglycemic coma due to diabetes mellitus | 421725003 | SNOMED | NO | YES | NO |
| 4228112 | Hypoglycemic coma due to type 1 diabetes mellitus | 421437000 | SNOMED | YES | YES | NO |
| 36714116 | Hypoglycemic coma due to type 2 diabetes mellitus | 719216001 | SNOMED | NO | YES | NO |
| 24609 | Hypoglycemia | 302866003 | SNOMED | NO | YES | NO |
| 23034 | Neonatal hypoglycemia | 52767006 | SNOMED | YES | YES | NO |
| 4029424 | Non-diabetic hypoglycemia | 237637005 | SNOMED | YES | YES | NO |
| 4029423 | Hypoglycemia due to diabetes mellitus | 237633009 | SNOMED | NO | YES | NO |
| 45769876 | Hypoglycemia due to type 1 diabetes mellitus | 84371000119108 | SNOMED | YES | YES | NO |
| 45757363 | Hypoglycemia due to type 2 diabetes mellitus | 120731000119103 | SNOMED | NO | YES | NO |
| 4096804 | Drug-induced hypoglycemia without coma | 190448007 | SNOMED | NO | YES | NO |

#### B.23 Hypotension

##### B.23.1 Cohort Entry Events

People enter the cohort when observing any of the following:

1. condition occurrences of ‘[LEGEND HTN] Hypotension’.

##### B.23.2 Cohort Exit

The cohort end date will be offset from index event’s start date plus 1 day.

##### B.23.3 Cohort Eras

Entry events will be combined into cohort eras if they are within 90 days of each other.

##### B.23.4 Concept: [LEGEND HTN] Hypotension

| Concept ID | Concept Name | Code | Vocabulary | Excluded | Descendants | Mapped |
| --- | --- | --- | --- | --- | --- | --- |
| 313232 | Hemodialysis-associated hypotension | 408667000 | SNOMED | YES | YES | NO |
| 317002 | Low blood pressure | 45007003 | SNOMED | NO | YES | NO |
| 314432 | Maternal hypotension syndrome | 88887003 | SNOMED | YES | YES | NO |

#### B.24 Joint pain

##### B.24.1 Cohort Entry Events

People enter the cohort when observing any of the following:

1. condition occurrences of ‘Joint pain’.

##### B.24.2 Cohort Exit

The cohort end date will be offset from index event’s start date plus 1 day.

##### B.24.3 Cohort Eras

Entry events will be combined into cohort eras if they are within 90 days of each other.

##### B.24.4 Concept: Joint pain

| Concept ID | Concept Name | Code | Vocabulary | Excluded | Descendants | Mapped |
| --- | --- | --- | --- | --- | --- | --- |
| 77074 | Joint pain | 57676002 | SNOMED | NO | NO | NO |

#### B.25 Lower extremity amputation

##### B.25.1 Cohort Entry Events

People may enter the cohort when observing any of the following:

1. procedure occurrences of ‘below-knee amputations’.

Restrict entry events to having no procedure occurrences of ‘below-knee amputations’, starting in the 30 days prior to cohort entry start date.

##### B.25.2 Cohort Exit

The cohort end date will be offset from index event’s start date plus 0 days.

##### B.25.3 Cohort Eras

Entry events will be combined into cohort eras if they are within 0 days of each other.

##### B.25.4 Concept: below-knee amputations

| Concept ID | Concept Name | Code | Vocabulary | Excluded | Descendants | Mapped |
| --- | --- | --- | --- | --- | --- | --- |
| 4264289 | Amputation of ankle | 397218006.00 | SNOMED | NO | YES | NO |
| 2006242 | Amputation of ankle through malleoli of tibia and fibula | 84.14 | ICD9Proc | NO | YES | NO |
| 2105446 | Amputation, leg, through tibia and fibula | 27880.00 | CPT4 | NO | YES | NO |
| 2105804 | Amputation, foot; midtarsal (eg, Chopart type procedure) | 28800.00 | CPT4 | NO | YES | NO |
| 2105805 | Amputation, foot; transmetatarsal | 28805.00 | CPT4 | NO | YES | NO |
| 2105806 | Amputation, metatarsal, with toe, single | 28810.00 | CPT4 | NO | YES | NO |
| 2105807 | Amputation, toe; metatarsophalangeal joint | 28820.00 | CPT4 | NO | YES | NO |
| 2105808 | Amputation, toe; interphalangeal joint | 28825.00 | CPT4 | NO | YES | NO |
| 2105451 | Amputation, ankle, through malleoli of tibia and fibula (eg, Syme, Pirogoff type procedures), with plastic closure and resection of nerves | 27888.00 | CPT4 | NO | YES | NO |
| 2105447 | Amputation, leg, through tibia and fibula; with immediate fitting technique including application of first cast | 27881.00 | CPT4 | NO | YES | NO |
| 4338257 | Amputation of leg through tibia and fibula | 88312006.00 | SNOMED | NO | YES | NO |
| 2105448 | Amputation, leg, through tibia and fibula; open, circular (guillotine) | 27882.00 | CPT4 | NO | YES | NO |
| 4108565 | Amputation of the foot | 180030006.00 | SNOMED | NO | YES | NO |
| 2006229 | Amputation of toe | 84.11 | ICD9Proc | NO | YES | NO |
| 4159766 | Amputation of toe | 371186005.00 | SNOMED | NO | YES | NO |
| 4054983 | Amputation through foot | 211570009.00 | SNOMED | NO | YES | NO |
| 2006230 | Amputation through foot | 84.12 | ICD9Proc | NO | YES | NO |
| 4143797 | Amputation through metatarsal bones | 265739006.00 | SNOMED | NO | YES | NO |
| 2105450 | Amputation, leg, through tibia and fibula; re-amputation | 27886.00 | CPT4 | NO | YES | NO |
| 2006231 | Disarticulation of ankle | 84.13 | ICD9Proc | NO | YES | NO |
| 2006244 | Disarticulation of knee | 84.16 | ICD9Proc | NO | YES | NO |
| 4018719 | Midtarsal amputation of foot | 209724005.00 | SNOMED | NO | YES | NO |
| 2006243 | Other amputation below knee | 84.15 | ICD9Proc | NO | YES | NO |
| 2105449 | Amputation, leg, through tibia and fibula; secondary closure or scar revision | 27884.00 | CPT4 | YES | YES | NO |
| 4219032 | Amputation of lower limb | 397117006.00 | SNOMED | NO | YES | NO |

#### B.26 Nausea

##### B.26.1 Cohort Entry Events

People enter the cohort when observing any of the following:

1. condition occurrences of ‘[LEGEND HTN] Nausea’.

##### B.26.2 Cohort Exit

The cohort end date will be offset from index event’s start date plus 1 day.

##### B.26.3 Cohort Eras

Entry events will be combined into cohort eras if they are within 30 days of each other.

##### B.26.4 Concept: [LEGEND HTN] Nausea

| Concept ID | Concept Name | Code | Vocabulary | Excluded | Descendants | Mapped |
| --- | --- | --- | --- | --- | --- | --- |
| 30284 | Motion sickness | 37031009 | SNOMED | YES | YES | NO |
| 31967 | Nausea | 422587007 | SNOMED | NO | YES | NO |

#### B.27 Peripheral edema

##### B.27.1 Cohort Entry Events

People enter the cohort when observing any of the following:

1. condition occurrences of ‘Edema’.

##### B.27.2 Cohort Exit

The cohort end date will be offset from index event’s start date plus 1 day.

##### B.27.3 Cohort Eras

Entry events will be combined into cohort eras if they are within 180 days of each other.

##### B.27.4 Concept: Edema

| Concept ID | Concept Name | Code | Vocabulary | Excluded | Descendants | Mapped |
| --- | --- | --- | --- | --- | --- | --- |
| 433595 | Edema | 267038008 | SNOMED | NO | YES | NO |
| 133299 | Swelling of limb | 80068009 | SNOMED | NO | YES | NO |

#### B.28 Photosensitivity

##### B.28.1 Cohort Entry Events

People enter the cohort when observing any of the following:

1. condition occurrences of ‘Photosensitivity’.

##### B.28.2 Cohort Exit

The cohort end date will be offset from index event’s start date plus 1 day.

##### B.28.3 Cohort Eras

Entry events will be combined into cohort eras if they are within 90 days of each other.

##### B.28.4 Concept: Photosensitivity

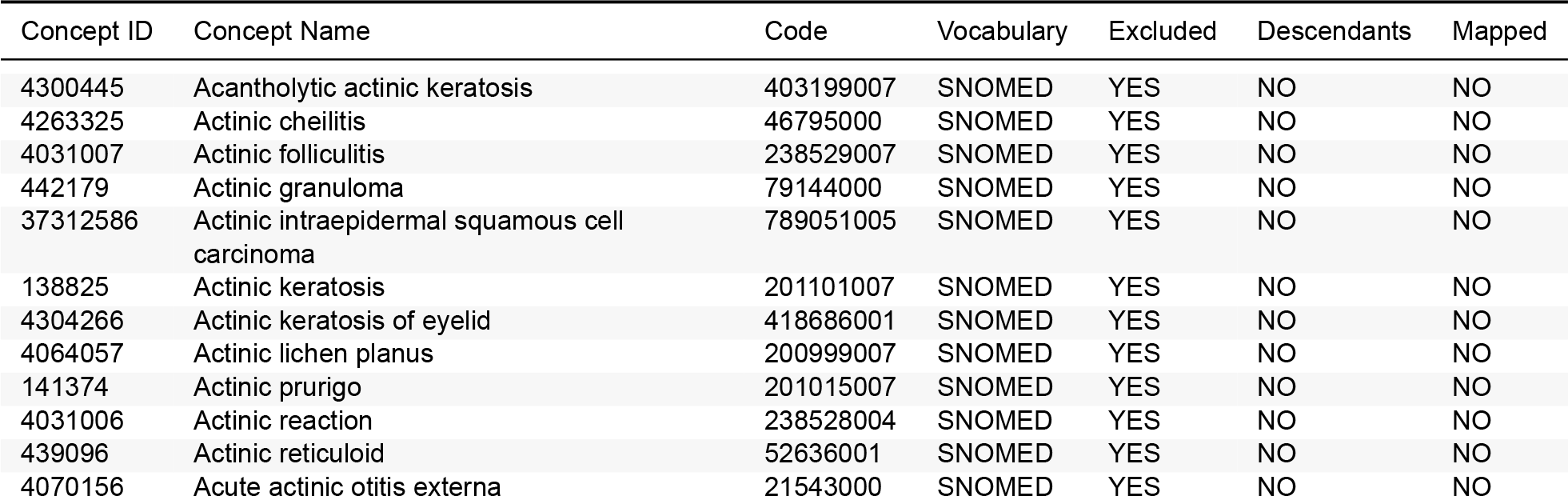
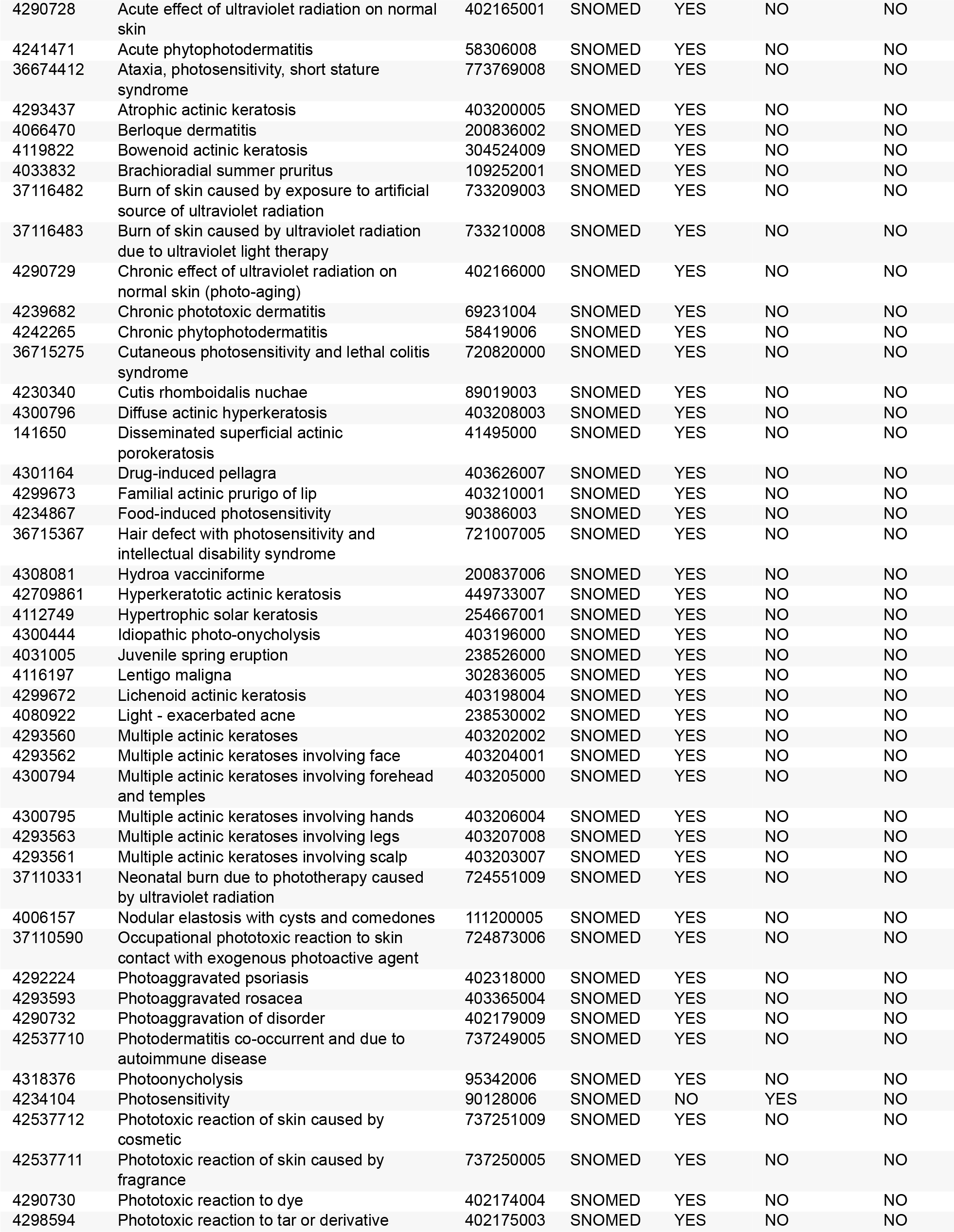
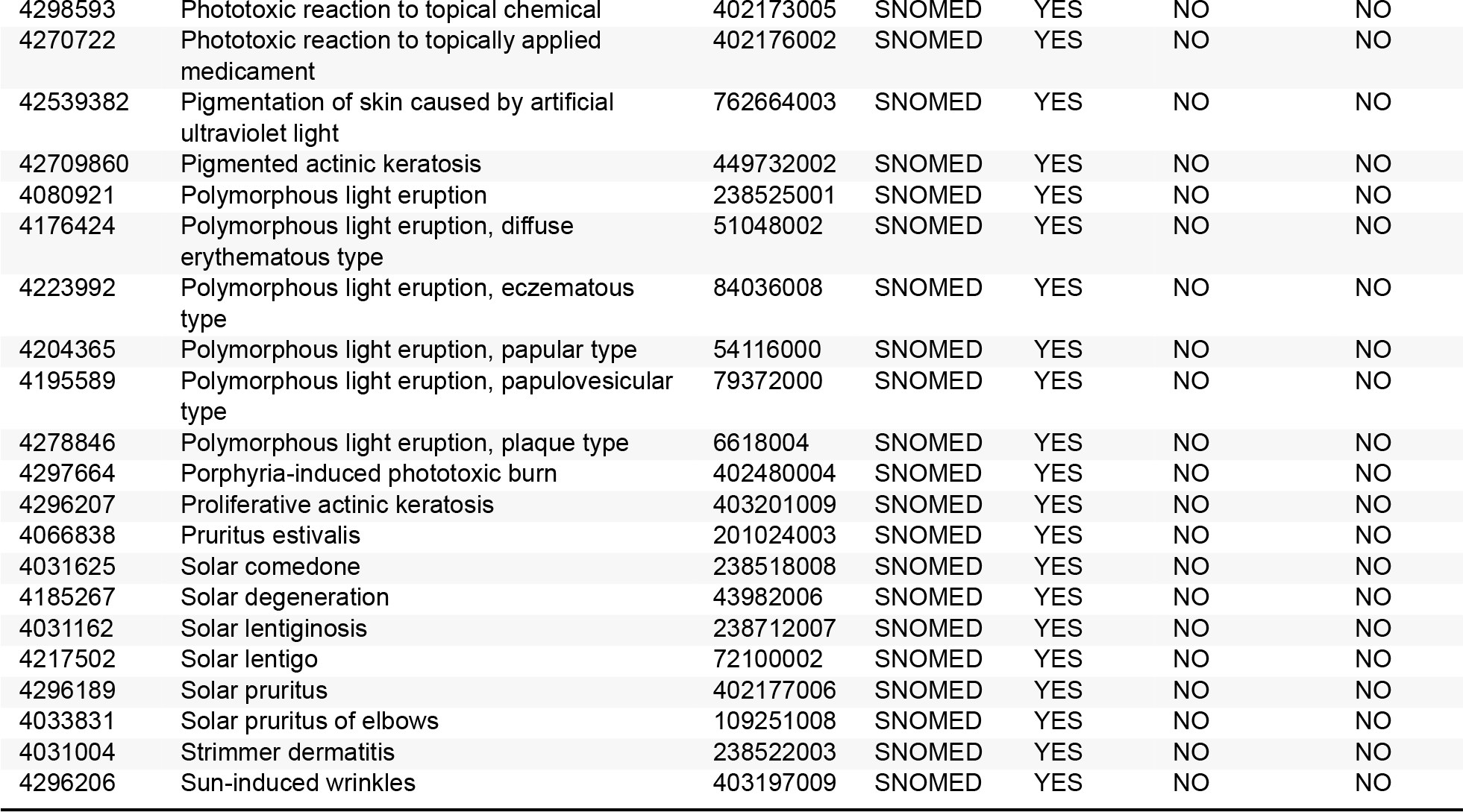

#### B.29 Renal cancer

##### B.29.1 Cohort Entry Events

People with continuous observation of 365 days before event enter the cohort when observing any of the following:

1. condition occurrence of ‘Primary malignant neoplasm of kidney’ for the first time in the person’s history.

Limit cohort entry events to the earliest event per person.

##### B.29.2 Cohort Exit

The person also exists the cohort at the end of continuous observation.

##### B.29.3 Cohort Eras

Entry events will be combined into cohort eras if they are within 0 days of each other.

##### B.29.4 Concept: Primary malignant neoplasm of kidney

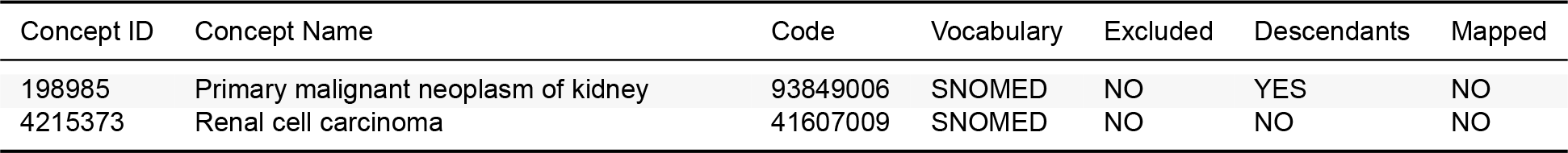

| Concept ID | Concept Name | Code | Vocabulary | Excluded | Descendants | Mapped |
| --- | --- | --- | --- | --- | --- | --- |
| 198985 | Primary malignant neoplasm of kidney | 93849006 | SNOMED | NO | YES | NO |
| 4215373 | Renal cell carcinoma | 41607009 | SNOMED | NO | NO | NO |

#### B.30 Thyroid tumor

##### B.30.1 Cohort Entry Events

People with continuous observation of 365 days before event enter the cohort when observing any of the following:

1. condition occurrence of ‘Neoplasm of thyroid gland’ for the first time in the person’s history.

Limit cohort entry events to the earliest event per person.

##### B.30.2 Cohort Exit

The person also exists the cohort at the end of continuous observation.

##### B.30.3 Cohort Eras

Entry events will be combined into cohort eras if they are within 0 days of each other.

##### B.30.4 Concept: Neoplasm of thyroid gland

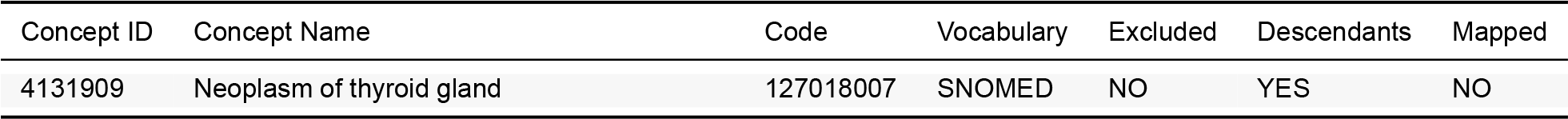

| Concept ID | Concept Name | Code | Vocabulary | Excluded | Descendants | Mapped |
| --- | --- | --- | --- | --- | --- | --- |
| 4131909 | Neoplasm of thyroid gland | 127018007 | SNOMED | NO | YES | NO |

#### B.31 Venous thromboembolism

##### B.31.1 Cohort Entry Events

People enter the cohort when observing any of the following:

1. condition occurrences of ‘[LEGEND HTN] Venous thromboembolism (pulmonary embolism and deep vein thrombosis)’.

##### B.31.2 Cohort Exit

The cohort end date will be offset from index event’s start date plus 1 day.

##### B.31.3 Cohort Eras

Entry events will be combined into cohort eras if they are within 180 days of each other.

##### B.31.4 Concept: [LEGEND HTN] Venous thromboembolism (pulmonary embolism and deep vein thrombosis)

| Concept ID | Concept Name | Code | Vocabulary | Excluded | Descendants | Mapped |
| --- | --- | --- | --- | --- | --- | --- |
| 435616 | Amniotic fluid embolism | 17263003.0 | SNOMED | YES | YES | NO |
| 435887 | Antepartum deep vein thrombosis | 49956009.0 | SNOMED | YES | YES | NO |
| 196715 | Budd-Chiari syndrome | 82385007.0 | SNOMED | YES | YES | NO |
| 4062269 | Cerebral venous thrombosis in pregnancy | 200259003.0 | SNOMED | YES | YES | NO |
| 442055 | Obstetric air pulmonary embolism | 200286003.0 | SNOMED | YES | YES | NO |
| 433832 | Obstetric blood-clot pulmonary embolism | 200299000.0 | SNOMED | YES | YES | NO |
| 435026 | Obstetric pulmonary embolism | 200284000.0 | SNOMED | YES | YES | NO |
| 440477 | Obstetric pyemic and septic pulmonary embolism | 267284008.0 | SNOMED | YES | YES | NO |
| 318137 | Phlebitis and thrombophlebitis of intracranial sinuses | 192753009.0 | SNOMED | YES | YES | NO |
| 199837 | Portal vein thrombosis | 17920008.0 | SNOMED | YES | YES | NO |
| 438820 | Postpartum deep phlebothrombosis | 56272000.0 | SNOMED | YES | YES | NO |
| 440417 | Pulmonary embolism | 59282003.0 | SNOMED | NO | YES | NO |
| 254662 | Pulmonary infarction | 64662007.0 | SNOMED | NO | YES | NO |
| 4235812 | Septic thrombophlebitis | 439731006.0 | SNOMED | YES | YES | NO |
| 195294 | Thrombosed hemorrhoids | 75955007.0 | SNOMED | YES | YES | NO |
| 4187790 | Thrombosis of retinal vein | 46085004.0 | SNOMED | YES | YES | NO |
| 444247 | Venous thrombosis | 111293003.0 | SNOMED | NO | YES | NO |
| 44834756 | Acute venous embolism and thrombosis of other specified veins | 453.8 | ICD9CM | NO | NO | NO |

#### B.32 Vomiting

##### B.32.1 Cohort Entry Events

People enter the cohort when observing any of the following:

1. condition occurrences of ‘[LEGEND HTN] Vomiting’.

##### B.32.2 Cohort Exit

The cohort end date will be offset from index event’s start date plus 1 day.

##### B.32.3 Cohort Eras

Entry events will be combined into cohort eras if they are within 30 days of each other.

##### B.32.4 Concept: [LEGEND HTN] Vomiting

| Concept ID | Concept Name | Code | Vocabulary | Excluded | Descendants | Mapped |
| --- | --- | --- | --- | --- | --- | --- |
| 40480290 | Hyperemesis | 444673007 | SNOMED | YES | YES | NO |
| 4216862 | Postoperative vomiting | 72245005 | SNOMED | YES | YES | NO |
| 441408 | Vomiting | 422400008 | SNOMED | NO | YES | NO |
| 440785 | Vomiting of pregnancy | 90325002 | SNOMED | YES | YES | NO |

### C Negative Control Concepts

Table 72: Negative outcome controls specified through condition occurrences that map to (a descendent of) the indicated concept ID

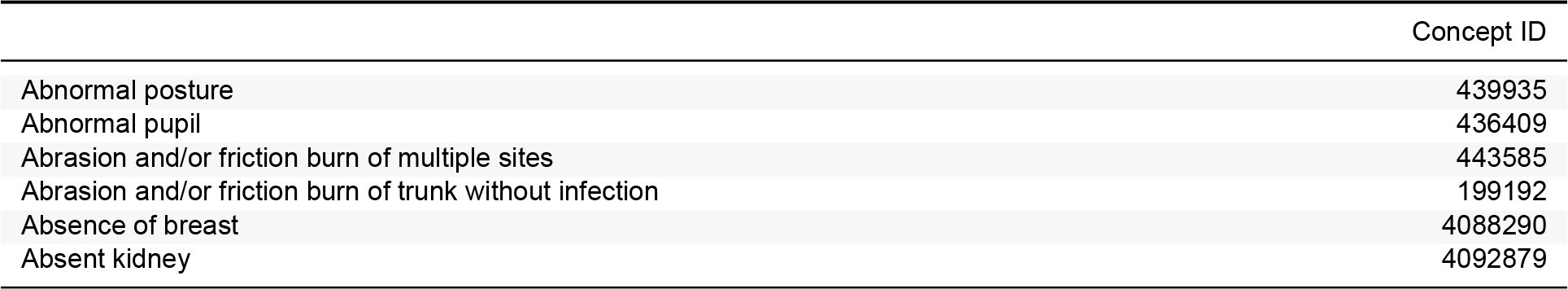
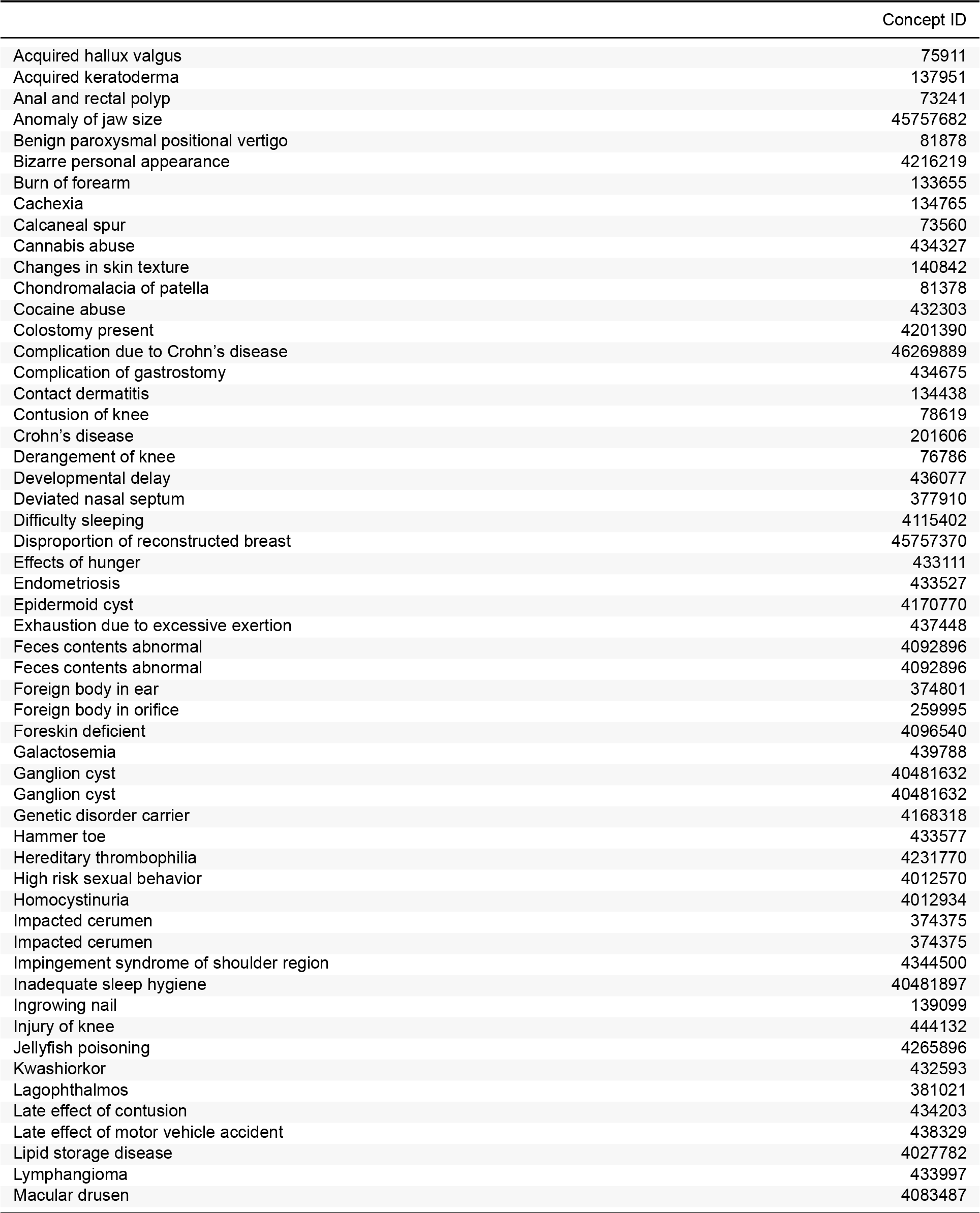
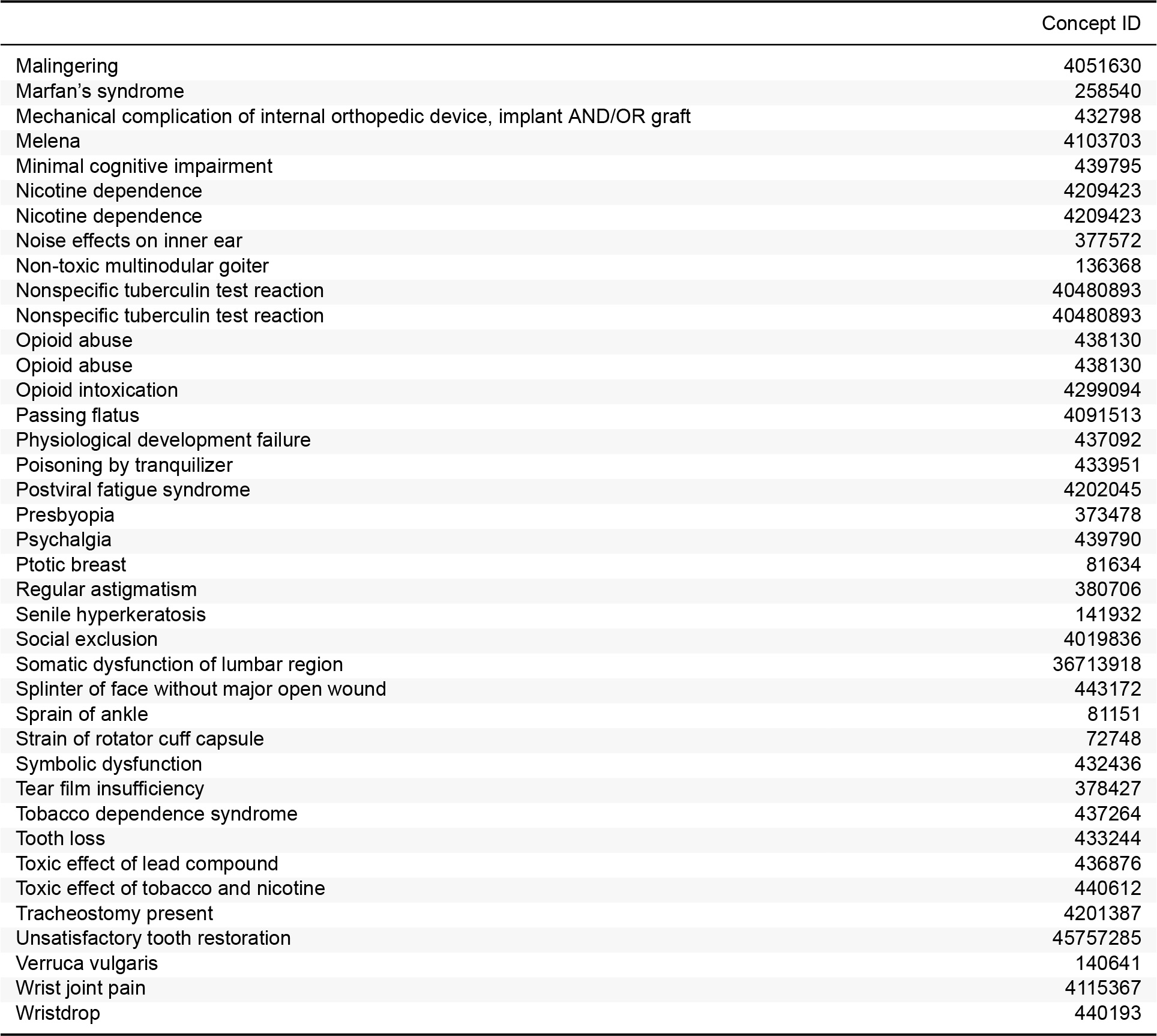

## Notes

### Funding Statement

The proposal was partially funded through the National Institutes of Health grants K23 HL153775 and R01 LM006910 and an Intergovernmental Personnel Act agreement with the US Department of Veterans Affairs. The funders had no role in the design and conduct of the proposal; preparation, review, or approval of the manuscript; and decision to submit the manuscript for publication.

### Author Declarations

IRB approval or waiver statements from partners are included in Table 4. The study uses a federated analytic model that is exempt from the purview of the IRB for data partners.

